# A Comparison of Manual and Automated Approaches to Developing Computable Algorithms for Identifying Acute Pancreatitis

**DOI:** 10.64898/2026.06.05.26354934

**Authors:** Maralyssa A Bann, David S Carrell, Susan Gruber, Patrick J Heagerty, Brian D Williamson, Jennifer C Nelson, Brian Hazlehurst, Andrew Felcher, Denis B Nyongesa, Matthew T Slaughter, Daniel S. Sapp, David J Cronkite, Robert Ball, James S. Floyd

## Abstract

**Objective:** Clinical phenotyping methods that rely on clinical and informatics expertise can be time-intensive and costly. We tested both manual and highly automated approaches using electronic health record (EHR) data to identify an FDA Sentinel Initiative health outcome of interest, acute pancreatitis.

**Materials and Methods:** We trained and evaluated machine learning algorithms using EHR data with two approaches: a custom approach that included manually curated features and trained on outcomes data validated with medical record review, and a highly automated approach that greatly simplifies and automates feature engineering and relies on low-cost silver-standard outcomes for model training.

**Results:** Custom algorithms using manually curated structured claims data discriminated cases from non-cases with a high degree of accuracy (cv-AUC 0.89 [95%CI 0.84-0.94]); the inclusion of natural language processing (NLP)-derived covariates from clinical notes increased performance slightly (cv-AUC 0.91[95%CI 0.86-0.97]). The automated algorithm trained on the outcome count of diagnosis codes performed less well (AUC 0.80 [95% CI 0.75-0.85]) but improved using maximum lipase value as an outcome (AUC 0.88 [95% CI 0.84-0.92]). At a positive predictive value of 90%, the custom algorithm had a sensitivity of 92%, the automated algorithm trained on diagnosis code count had a sensitivity of 45%, and the automated algorithm trained on maximum lipase value had a sensitivity of 84%. However, a prediction rule derived by clinicians during chart review was nearly as accurate (maximum lipase value ≥ 3 times upper limit of normal; AUC 0.86, PPV 85%, sensitivity 92%).

**Discussion:** Machine learning algorithms with manually curated structured data and NLP features trained on validated outcomes data successfully identified validated events. Use of an outcome in the automated model based on specific phenotype knowledge (maximum lipase value) allowed for performance similar to the custom model and with considerably less resources.

## BACKGROUND

Acute pancreatitis is a gastrointestinal illness characterized by inflammation of the pancreas that can cause abdominal pain, nausea, and vomiting. While most cases are mild and self-limited[1], around 20% may progress to severe complications such as pancreatic necrosis, pseudocyst formation, local or systemic vascular dysfunction, or persistent organ failure.[2,3] Severe disease is associated with a mortality rate of nearly 40%.[4] The incidence of acute pancreatitis is increasing[5] and it is a major cause of gastrointestinal-disease related hospital admissions.[6,7] Drug-induced acute pancreatitis may represent up to 5% of all cases and over 120 medications have been implicated as potential causative agents.[8] Methods to study acute pancreatitis as a medication-related adverse event using health care claims data have been limited by poor algorithm performance for identifying this outcome based on diagnosis codes.[9,10] Leveraging rich information from electronic health record (EHR) data including text from clinical documentation together with machine learning methods may improve the identification of acute pancreatitis and enhance ongoing drug safety monitoring efforts.

The United States Food and Drug Administration (FDA) began the Sentinel Initiative in 2008 to prospectively monitor real-world data for the safety of marketed medical products.[11] The resulting multisite distributed database of health care claims-based information and analytic tools to query it[12,13] allow for powerful safety surveillance methods such as the semi-automated Active Risk Identification and Analysis (ARIA) System.[14] While ARIA is currently in use to inform FDA decision-making, several health outcomes with complex clinical presentation remain a challenge due to inaccuracy of coded claims data for use as clinical outcomes for safety surveillance.[15, 16] For example, a diagnosis code for anaphylaxis has a positive predictive value (PPV) of only about 65%.[17–19] We recently demonstrated that application of machine learning methods and natural language processing (NLP) to rich clinical data in the EHR can improve identification of anaphylaxis to an extent that a computable outcome may be suitable for drug safety studies.[20] It is unknown whether the strategy of leveraging EHR works well for other acute clinical phenotypes, including acute pancreatitis – and if so, whether alternatives that incorporate more time and resource-efficient approaches may be sufficient.

Algorithm development relying on expert curation and manual annotation of gold-standard training sets requires considerable clinical and informatics expertise, making it time-intensive and costly; computable phenotyping approaches that address this inefficiency are needed.[21, 22] In the last few years, automated development of clinical phenotype algorithms has shown promise for several chronic health conditions.[23–28] PheNorm, a specific automated approach initially developed for the identification of chronic health conditions, is particularly appealing because it does not require costly and operator-dependent manual feature engineering or validated outcomes during the training phase but instead automates the feature engineering process and uses “silver-standard” labels, such as patient-level counts of diagnosis codes or NLP mentions of the outcome, for model training.[27] (Note: Evaluation of all models, whether developed by custom or automated approaches, requires some manually validated outcome data.) Automated approaches have rarely been applied to an acute health condition.[29] A better understanding of the relative performance of different clinical phenotyping approaches is an important goal toward advancing the scope of safety surveillance activities.

Therefore, the aim of this investigation is to investigate two alternative approaches to developing computable algorithms that use EHR data to identify episodes of acute pancreatitis: 1) a “custom” modeling approach with manually curated structured and NLP features trained on expert-validated outcomes, and 2) a highly automated modeling approach that identifies candidate features by automated text-mining of clinical knowledge articles and trains models on easily-operationalized silver-standard outcomes data. For simplicity throughout the text, we refer to these as “custom” and “automated” approaches, respectively.

## METHODS

### Setting and study population

This study used EHR data from Kaiser Permanente Northwest (KPNW), which delivers care to roughly 600,000 people in northwest Oregon and southwest Washington state. KPNW provides predominantly (> 95%) integrated care (HMO) insurance plans to its members and includes outpatient, emergency department (ED), and inpatient care.

Eligible KPNW members were 18 years and older and continuously enrolled for at least one year before an inpatient, ED, or outpatient encounter at a KPNW-owned facility with a diagnosis code for acute pancreatitis (K85) that occurred between October 1, 2015, and December 31, 2019 (a qualifying encounter). To identify incident (first) events, we excluded individuals with a code for acute (ICD-9 577.0; ICD-10 K85) or chronic pancreatitis (ICD-9 577.1; ICD-10 K86.0, K86.1) in the year before the first qualifying encounter. For individuals with more than one potential acute pancreatitis event during the study period, only the first event was eligible for sampling.

Because the treatment of acute pancreatitis often involves care in multiple settings, each potential event was classified according to the highest level of care with an acute pancreatitis code occurring within 7 days of the initial qualifying encounter. Thus, potential events that initially presented in the outpatient or ED setting could ultimately be classified as inpatient events.

Out of 1,843 potential acute pancreatitis events that met inclusion criteria, we randomly sampled 299 for validation with medical record review to create gold-standard outcome labels. The 299 observations that underwent medical record review were used in training and evaluating the algorithm developed by the custom approach, and in evaluating the algorithm developed by the automated approach. The 1,544 potential events that did not undergo medical record review were used to train our automated algorithms using silver-standard labels (which were then evaluated using the 299 gold standard outcome observations).

### Custom modeling approach

The custom model used:

- Physician supervised validation of acute pancreatitis events with medical record review to create gold-standard labels
- Manual curation of structured health care data covariates
- Manual curation of NLP-derived unstructured EHR text covariates
- Machine learning statistical methods (a large library of candidate algorithms including the Super Learner) using gold-standard training data to generate predictive algorithms
- Cross-validation to evaluate performance of each algorithm

#### Gold standard label creation

For the randomly chosen subset of 299 observations, we performed medical record reviews to determine whether potential events met clinical criteria for acute pancreatitis as defined by the revised Atlanta Classification, which requires the presence of at least two of the following: characteristic abdominal pain, elevated serum amylase or lipase laboratory values, or imaging findings consistent with pancreatic inflammation or its sequelae.[30] Two abstractors trained by physicians collected data and performed initial validation, followed by physician adjudication for any uncertain cases. In our recently published validation study that describes these methods [9], we found that the positive predictive value (PPV) for an acute pancreatitis diagnosis code was 61% (95% CI 55%-66%), and the additional presence of lipase laboratory value ≥ 3 times upper limit of normal resulted in a PPV of 92% (95%CI 86%-95%). Our expert clinicians corroborated the predictive utility of lipase laboratory values for acute pancreatitis in their own medical decision-making and the common use of this quantitative threshold. We therefore incorporated this knowledge into the custom modeling approach through manual curation of covariates and into the automated modeling approach through creation of a corresponding silver-standard outcome (maximum lipase laboratory value). We also used the findings from this validation study as comparison for custom and automated modeling approaches.

#### Manual curation of features

Guided by clinical domain expertise and informatics knowledge, we selected 122 claims-based structured predictors (Supplemental Materials Appendix A). Illustrative examples of these covariates include patient demographics, setting of care, diagnosis codes for acute pancreatitis causes/clinical risk factors (e.g., hypertriglyceridemia, alcohol use disorder, gallbladder/biliary disease, etc.) or competing diagnoses (e.g., peptic ulcer disease, appendicitis, pancreatic cancer, etc.), procedure codes for abdominal imaging, and lipase labs. We used the Sentinel Common Data Model[31] and the HMO Research Network Virtual Data Warehouse[32] for standardized representations of these variables. Our clinical and informatics experts manually curated an initial custom dictionary of medical concepts and other NLP-derived features they considered useful for identifying acute pancreatitis events. This set of manually curated NLP-derived covariates (Supplemental Materials Appendix B) contained 266 candidate measures, including mentions of pain along with location (e.g., epigastric, abdominal, chest), nature, (e.g., radiating to back) or time course (e.g., recent onset, acute, chronic); other symptoms (e.g., nausea, vomiting); mentions of imaging findings (e.g., necrosis, fluid, or pseudocyst); radiologic conclusion language with the term “pancreatitis” (e.g., “consistent with” or “suggestive of” pancreatitis); competing diagnoses; and counts of affirmative mentions of pancreatitis.

#### Custom model development

Similar to our previous effort to model anaphylaxis, we used the Super Learner R package (v2.0-28) to consider multiple learning algorithms for prediction.[33] A preliminary dimension reduction step excluded covariates with low variation that were uncorrelated with the outcome (≥98% of observations having the same value and absolute correlation <0.1), and all but one of any set of highly correlated covariates (absolute correlation > 0.98). The data made available to the Super Learner (Supplemental Materials Appendix C) contained 299 observations (181 cases, 118 non-cases) and 220 covariates (59 structured, 161 NLP-derived). Following guidelines suggested by Phillips et al[34] we specified the area under the receiver-operating characteristic curve (AUC) loss function, a 20-fold cross validation scheme where assignment to the disjoint validation sets was stratified by the outcome to ensure that the distribution of the outcome in each corresponding (independent) training set matched the overall prevalence. The Super Learner library consisted of eight parametric and machine learning algorithms [logistic regression (GLM), elastic net (eNet), two variants of gradient boosting (XGboost), two variants of bayesian additive regression trees (BART) and two neural net architectures (NNET)] coupled with three covariate retention strategies:[35–41]

1. Retain all covariates
2. PAM: partitioning around medoids, choosing to retain between 30 and 100 covariates, based on optimizing the silhouette width
3. Lasso: retain a minimum of 10 covariates with non-zero coefficients in a lasso model based on their conditional associations with the outcome

To investigate the impact of incorporating NLP-derived covariates we first allowed the Super Learner to consider only the 59 structured covariates, and then supplied it with both structured and NLP covariates.

We evaluated the performance of these models based on cross-validated AUC. For the best performing models, we also calculated cross-validated sensitivity, specificity, PPV, NPV, and F1 score (the harmonic mean of PPV and sensitivity). To identify important predictors in the model we ranked them by variable importance defined as the marginal mean difference in predicted probability associated with a 1-unit change in a binary covariate or a 1-standard-deviation change in a nonbinary covariate.

### Automated modeling approach

The automated model used:

- Easily operationalized surrogates for acute pancreatitis to create silver-standard labels
- Automated curation of NLP-derived covariates
- A limited number of easily operationalized structured data covariates (e.g., age, sex)
- Machine learning statistical methods (linear regression with final outcome predictions based on a normal mixture model) using silver-standard training data to generate predictive algorithms
- Independent validation set to evaluate performance of each algorithm

We implemented approaches described by Yu, et al.[27] and further elaborated by Zhang et al.[28] to automate feature engineering using text mining of clinical knowledge articles and to train models based on silver standard outcome labels (imperfect predictors of the health outcome of interest) instead of validated outcomes data.

#### Silver standard label creation

We defined four silver standard outcome labels:

- Count of encounters with acute pancreatitis diagnosis codes
- Count of distinct affirmative mentions of “acute pancreatitis” or its synonyms appearing in clinical notes
- The sum of the first two silver labels (encounters and affirmative mentions)
- The highest lipase laboratory value within +/-14 days of the patient’s index date

Counts include all data for a given patient during the catchment period. Our first three silver labels are examples of data-driven measures that can be easily computed for each patient, can be curated without expert knowledge, and are believed to be correlated with the phenotype (following the method described by Yu and colleagues). Our fourth label, lipase lab value, is an example of a silver label based on clinical expert judgment and phenotype domain knowledge; Zhang and colleagues recommend using such labels if they have solid clinical rationale and are relatively easy to operationalize.

Automated feature engineering requires specification of a data catchment period, the window of time EHR data is used to generate patient-specific NLP-derived features and silver labels. In contrast to a years-long catchment period that might be appropriate for chronic conditions, we used a data catchment period of 14 days before to 30 days after date of first acute pancreatitis diagnosis code for clinical notes; 7 days before to 7 days after for imaging reports; and 14 days before to 14 days after for lipase tests. Our study corpus included all structured claims data, clinical encounter notes from the EHR, and imaging report notes from the EHR during the data catchment period. Patient after-visit summaries and patient instructions were excluded because these documents are likely to involve hypothetical mentions of medical concepts, making them poor indicators of phenotype status.

#### Automated NLP-derived covariates

Following the “concept collection” step of the Automated Feature Extraction for Phenotyping (AFEP) method,[24] we used MetaMap Lite 3.6.2rc5[42,43] and the UMLS 2020AB 0+4+9 vocabularies (built for MetaMap Lite and included on the same page) to identify medical concepts relevant to acute pancreatitis that appeared in ≥ 3 of 5 published acute pancreatitis clinical knowledge base articles.[44–48] Although use of expert review to remove concepts that are non-specific to the condition of interest is permitted by AFEP, we refrained from doing so in order to maintain a strictly automated curation process. The resulting dictionary included 167 unique concepts (Supplemental Materials Appendix D). We identified all affirmative instances of these concepts in patients’ clinical notes to create 167 corresponding count features for each potential event. In addition, we created 167 additional features by normalizing the original 167 counts by the character count of each patient’s clinical text.

#### Automated model development

Following the approach of Yu and colleagues, we trained one model for each of the four silver labels using data from each of the 1,544 potential events in our training sample. Candidate predictors included the 334 automated NLP-derived features (nonnormalized and normalized versions of the 167 count features described above), the three silver labels not used as the model’s silver outcome label, and our structured data measures for encounter days by care setting (ambulatory/outpatient, emergency department, or inpatient), age, sex, race, and Hispanic ethnicity. In a preliminary dimension reduction step,[26] we excluded NLP features that had rank correlation with the UMLS concept for acute pancreatitis less than 0.15; after this dimension reduction step, 125 NLP features remained.

For each model, one per silver label, the silver label is transformed to a normal mixture distribution and then self-regressed with dropout to denoise based on additional candidate features. Each model uses linear regression to relate the outcome (the given silver label) and candidate features. As recommended, we averaged the predicted probabilities from each individual model to create a fifth aggregate set of predicted probabilities.

We evaluated the performance of all models based on AUC estimated using the 299 observations with gold standard labels that were not used for training (an independent validation set). We also calculated sensitivity, specificity, PPV, NPV, and F1 score.

This Sentinel activity is a public health surveillance activity conducted under the authority of the FDA, and, accordingly, was not subject to institutional review board oversight.[49–51] All analyses were run in R (v3.6.3; R Core Team, 2020).

## RESULTS

### Custom modeling

Using structured claims data alone, estimates of the cross-validated AUC (cv-AUC) and 95% confidence intervals show that many algorithms achieved a cv-AUC of 0.89 or 0.90, indicating excellent ability to distinguish between cases and non-cases (Figure 1a, Supplemental Materials Appendix E Table 1). BART coupled with any of the retention strategies and NNET coupled with the Lasso retention strategies had the highest cv-AUCs (0.90), suggesting that the optimal prediction function contains some non-linearities and/or interaction terms not included in the main terms of the regression models. However, parsimony and interpretability favor the elastic net (Enet) coupled with the Lasso pre-screener, which had a similar cv-AUC of 0.89 (95% CI 0.84-0.94).

Including manually curated NLP-derived covariates led to small increases in cv-AUCs, although high dimensionality caused some of the neural net-based algorithms to fail. (Figure 1b, Supplemental Materials Appendix E Table 2). Super Learner, BART1-RetainAll, and BART2-Lasso had cv-AUCs of 0.93 (95% CI 0.89-0.98), while Enet-Lasso had a cv-AUC of 0.91 (95% CI 0.86-0.97). Coefficients for retained covariates and variable importance rankings for the Enet-Lasso models are displayed in Supplemental Materials Appendix F Tables 1-4. In both models, “lipase level greater than 3 times the upper limit of normal” was the most important predictor.

**Figure 1.**
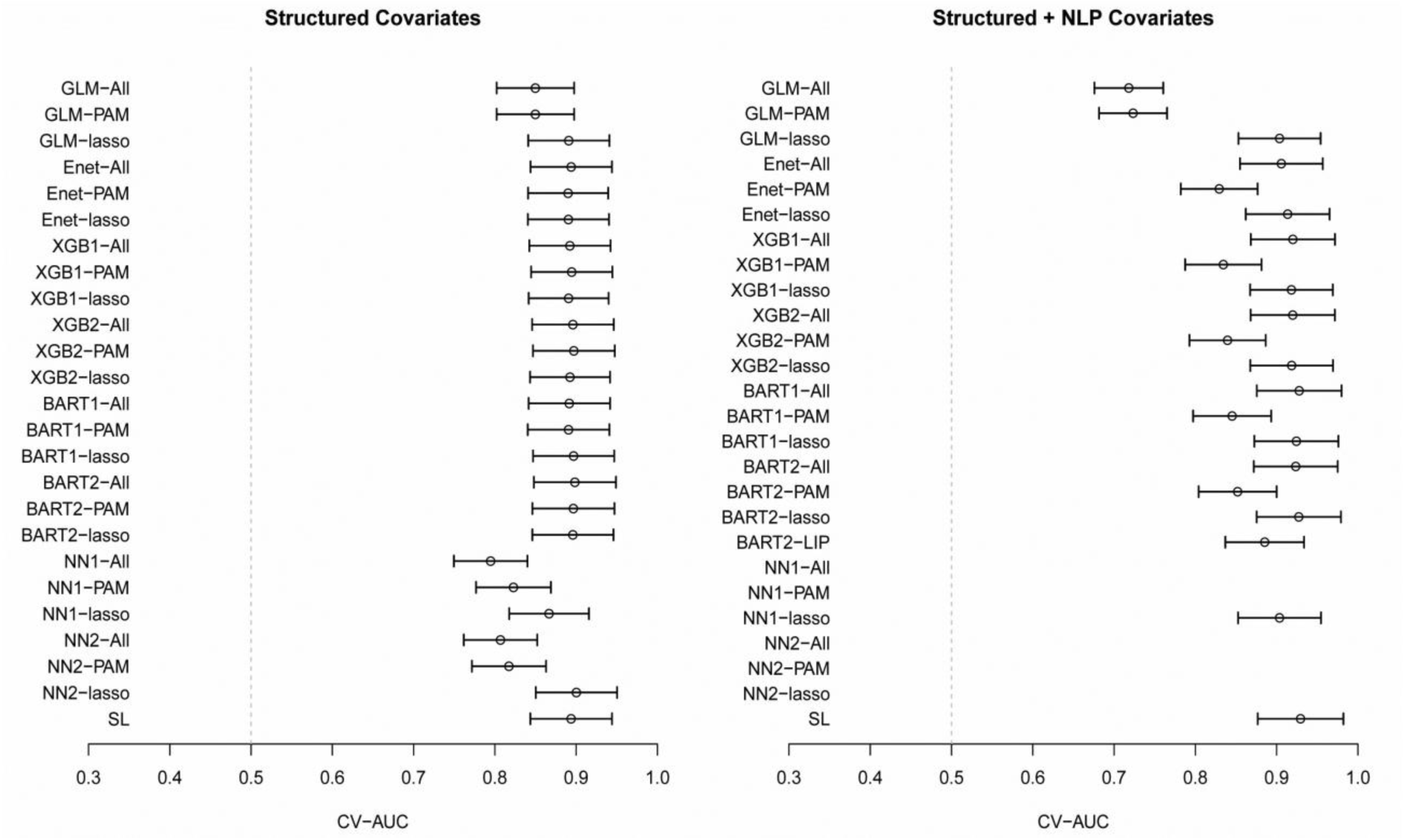
Cross-validated AUC and 95% confidence interval for each custom algorithm applied to manually curated structured covariates alone (1a, left) and to manually curated structured and NLP-derived covariates (1b, right)

For the Enet-Lasso model, we evaluated cross-validated sensitivity, specificity, positive predictive value (cv-PPV) and negative predictive value (cv-NPV). When only structured data features were included, 11 covariates were retained. At a classification threshold at the 40^th^ percentile of the predicted probabilities, the cv-PPV was 0.90 and cv-sensitivity was 0.88 (Figure 2a). When both structured data and NLP features were included, 13 covariates were retained. At a classification threshold at the 39^th^ percentile of predicted probabilities, the cv-PPV was 0.90 and cv-sensitivity was 0.92 (Figure 2b). Overall, the addition of NLP-derived features appeared to add little to the accuracy of the prediction models.

**Figure 2.**
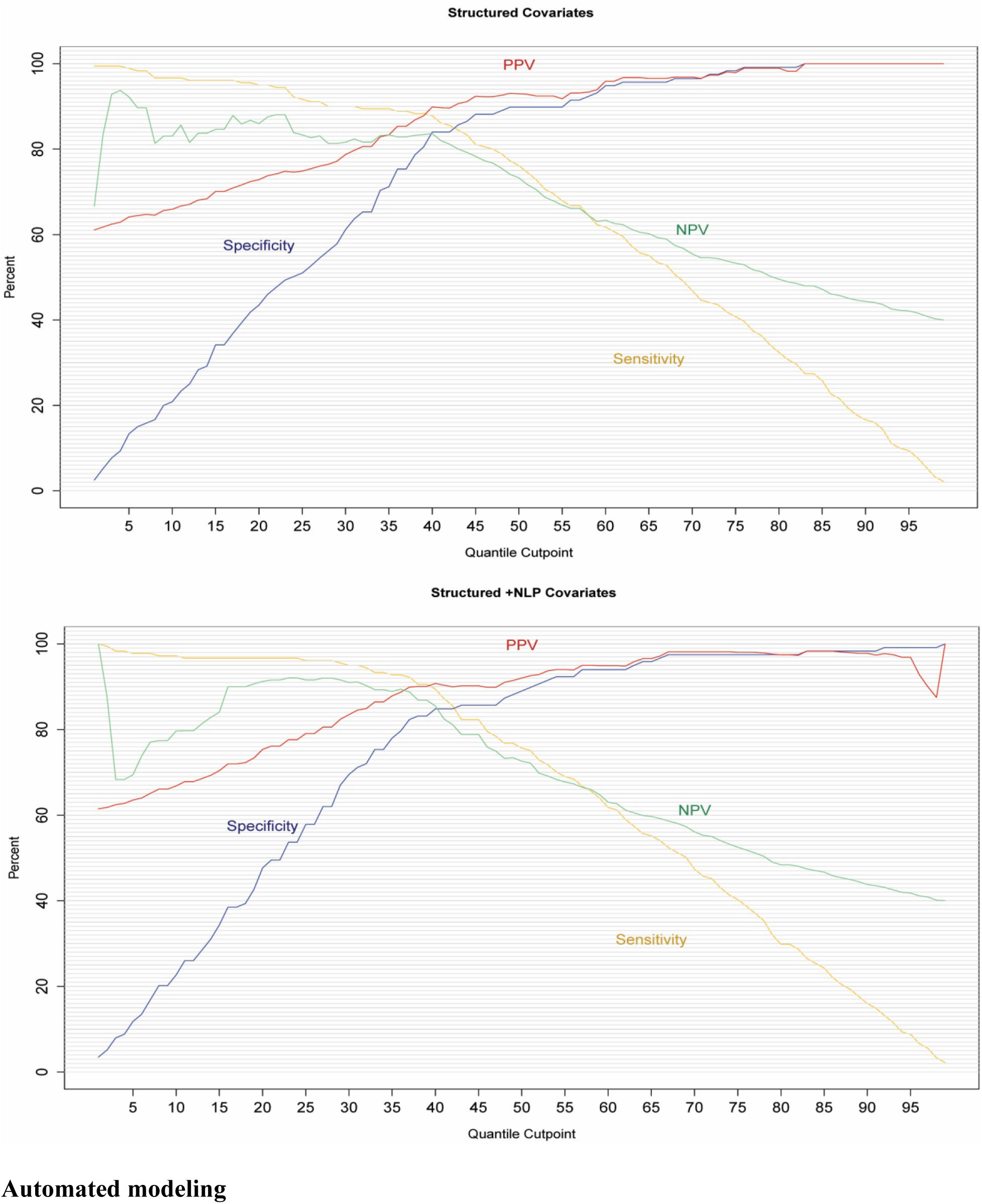
Cross-validated sensitivity, specificity, PPV, NPV for custom Enet-Lasso model trained on manually curated structured covariates alone (2a, top) and on manually curated structured and NLP-derived covariates (2b, bottom)

### Automated modeling

Compared to the custom models developed using manually curated features, the models using automated curation of features with silver standard outcomes of acute pancreatitis diagnosis codes or NLP mentions did not perform as well (Table 1).

**Table 1.** Performance of acute pancreatitis predictive modeling approaches.

|  |  | Fixing PPV at 90% |  |  |  |
| --- | --- | --- | --- | --- | --- |
|  | AUC (95% CI)* | PPV | Sens | Max F1 | Quantile |
| Chart review-derived predictive rule (max lipase $\geq 3$x ULN) | 0.86 | 0.85 | 0.92 | -- | -- |
| Custom/manually curated models |  |  |  |  |  |
| Structured covariates only (Enet-Lasso) | 0.89 (0.84-0.94) | 0.90 | 0.88 | 0.88 | 0.40 |
| Structured plus NLP covariates (Enet-Lasso) | 0.91 (0.86-0.97) | 0.90 | 0.92 | 0.91 | 0.37 |
| Automated models |  |  |  |  |  |
| AP diagnosis codes | 0.80 (0.75-0.85) | 0.90 | 0.45 | 0.60 | 0.69 |
| NLP mentions of AP | 0.72 (0.66-0.78) | 0.90 | 0.22 | 0.36 | 0.85 |
| Sum of diagnosis codes and NLP | 0.74 (0.69-0.80) | 0.90 | 0.30 | 0.90 | 0.79 |
| Maximum lipase level | 0.88 (0.84-0.92) | 0.90 | 0.84 | 0.87 | 0.42 |
| Aggregate | 0.85 (0.80-0.89) | 0.90 | 0.43 | 0.59 | 0.70 |

*\*As described in the text, manually curated models were evaluated with cv-AUC and automated models were evaluated using AUC*

For the automated model using silver standard outcome of counts of diagnosis codes, the AUC was 0.80 (95%CI 0.75-0.85). At a PPV of 90% the sensitivity was only 45%, markedly lower than the sensitivity of 92% from the manually curated model and also lower than the sensitivity of 92% from the chart review-derived prediction rule of maximum lipase greater than or equal to three times upper limit of normal.[9] When maximum lipase level was used as the silver standard outcome, the performance of the automated model approached that of the best manually curated models with AUC of 0.88 (95%CI 0.84-0.92). We also examined the predictive performance of the silver standard labels themselves, without any of the structured or NLP features. The AUCs for diagnosis code counts, NLP mention counts, sum of diagnosis code and NLP mentions, and maximum lipase level were 0.74 (95%CI 0.66-0.82), 0.70 (95%CI 0.64-0.77), 0.73 (95%CI 0.67-0.80), and 0.87 (95%CI 0.83-0.91) respectively.

## DISCUSSION

In this study, we found that a custom approach to algorithm development with manual curation of structured data features trained on validated outcomes using machine learning statistical methods achieved a high level of performance, and that the addition of NLP-derived features from unstructured clinical text improved performance only slightly. These custom models outperformed the three standard automated models (using diagnosis counts, NLP mentions, or the sum of the two as silver standard labels). Fixing PPV at 90%, the best manual custom model had a sensitivity of 92% while the best standard automated model had a sensitivity of 45%. The automated model trained on the silver standard outcome label of maximum lipase value performed nearly as well as the custom model, which is notable because the automated approach requires considerably fewer resources and less time to execute. The success of this silver standard outcome is likely due to the high correlation of lipase levels with validated acute pancreatitis and corroborates the use of domain knowledge to select appropriate silver standard outcomes. Neither the custom model with and without inclusion of NLP-derived predictors nor the automated approaches resulted in a meaningful improvement over a simple predictor based on maximum lipase value ≥ 3 times upper limit of normal, which was identified during our previous validation study (PPV 85%, sensitivity 92%).

This study is one of the first reports of the application of an automated modeling approach to an acute clinical phenotype, and in particular, is unique in its application to an FDA drug safety health outcome of interest. In contrast with the original publication describing PheNorm,[27] which reported cv-AUCs ranging from 0.90 to 0.95 when using patient-level counts of diagnosis codes and/or NLP mentions of the phenotype as silver standard labels for the chronic health conditions being modeled (such as coronary artery disease and rheumatoid arthritis), we found that the best-performing of these standard silver standard labels resulted in an AUC of only 0.80 for acute pancreatitis, translating to a sensitivity of less than 50% at a PPV of 90%. Another published report of PheNorm applied to a chronic (rheumatoid arthritis) and a subacute disease (active pulmonary tuberculosis, which requires treatment for several months) resulted in cvAUCs of 0.94 and 0.99 respectively.[52] One explanation for this discrepancy is that a count of healthcare encounters with a diagnoses code for a disease is likely to be a reliable indicator of the probability of that disease for conditions that require management over a long period of time. For acute events, this assumption may be unlikely to hold. Indeed, severe acute pancreatitis events that result in hospitalization directly from an emergency department visit may result in single healthcare encounter with a qualifying acute pancreatitis diagnosis code.

PheNorm was clearly designed to be a general informatics framework for automation of phenotyping in electronic health data. The primary features of this framework are automating the engineering of features extracted from clinical note text and use of easily implemented “silver standard” outcome labels in place of costly gold standard labels for model training. The PheNorm framework [27] and its subsequent elaborations [28] address many of the bottlenecks in electronic phenotyping while leaving open many possible modifications and improvements.

Future automated modeling efforts should explore incorporating other machine learning models, [53] novel approaches to defining silver standard labels optimized for acute health conditions, and augmenting silver standard labels with a limited amount of gold standard label during model training. In our study of acute pancreatitis, we added a silver label (maximum lipase lab level) based on clinical domain knowledge, an idea that fits within the PheNorm framework[28].

The original PheNorm report did not investigate whether automated creation and selection of NLP features improved the prediction of validated outcomes beyond the silver standard labels themselves, which may have been highly predictive of the presence of the chronic disease studied. In our study, we found that the structured and NLP features added little to the prediction of validated outcomes, except for the silver standard label diagnosis code counts (AUC 0.74 to 0.80). This finding suggests that future studies of automated modeling implementation should establish the suitability of the silver standard labels for their purpose; if the silver standard labels are strong but imperfect surrogates for the validated outcomes, it is unclear whether the addition of NLP features will meaningfully improve performance.

Our study had several strengths, including the availability of comprehensive EHR data from an integrated healthcare system and a set of gold standard labeled cases that were validated using established clinical criteria. There are also important limitations. Our study was conducted in a single healthcare system, and variation in documentation, billing, and coding practices may limit generalizability to other settings. Much of the diagnosis of acute pancreatitis is based on a widely available laboratory test that is well captured by EHR data. Similarly, a machine learning algorithm for rhabdomyolysis defined by laboratory test results improved on human-expert derived algorithms using only claims-based predictors.[54] More complex conditions for which diagnosis depends on constellations of signs and symptoms, such as anaphylaxis, may yield different findings. Additional research on the application of PheNorm and other automated modeling approaches to acute diseases is urgently needed to determine whether this approach holds promise as way to conduct efficient and timely drug safety studies.

## Data Availability

Data beyond what is contained in the manuscript will not be available to external sources.

## Acknowledgements

We would like to acknowledge the contributions of other members of the Sentinel Acute Pancreatitis Project whose input and assistance greatly contributed to the success of this work: Adebola Ajao, Kara Cushing-Haugen, Vina Graham, Luesa Healy, Sara Karami, Yong Ma, Kim Olson, Xu Shi, Mayura Shinde.

The views expressed in this paper represent those of the authors and do not necessarily represent the official views of the U.S. FDA.

This project was supported by Task Order 75F40119F19002 under Master Agreement 75F40119D10037 from the U.S. Food and Drug Administration (FDA). The FDA approved the study protocol including statistical analysis plan and reviewed and approved this manuscript.

Coauthors from the FDA participated in the results interpretation and in the preparation and decision to submit the manuscript for publication. The FDA had no role in data collection, management, or analysis.

