## Supplemental Materials for "A Comparison of Manual and Automated Approaches to Developing Computable Algorithms for Identifying Acute Pancreatitis"

**Supplementary Appendices**

### Appendix A: Manually Curated Structured Data Covariates

In collaboration between clinicians and informaticists, we manually engineered the following set of structured data covariates.

| Covariate Name | Description |
|---|---|
| AGE | Age in years at encounter |
| K85_1 | Diagnoses recorded for the qualifying encounter on EVENT_DATE included ICD-10 code K85.1, Biliary acute pancreatitis<br>N=No; Y=Yes |
| K85_10 | Diagnoses recorded for the qualifying encounter on EVENT_DATE included ICD-10 code K85.10, Biliary acute pancreatitis without necrosis or infection<br>N=No; Y=Yes |
| K85_2 | Diagnoses recorded for the qualifying encounter on EVENT_DATE included ICD-10 code K85.2, Alcohol induced acute pancreatitis<br>N=No; Y=Yes |
| K85_20 | Diagnoses recorded for the qualifying encounter on EVENT_DATE included ICD-10 code K85.20, Alcohol induced acute pancreatitis without necrosis or infection<br>N=No; Y=Yes |
| K85_3 | Diagnoses recorded for the qualifying encounter on EVENT_DATE included ICD-10 code K85.3, Drug induced acute pancreatitis<br>N=No; Y=Yes |
| K85_8 | Diagnoses recorded for the qualifying encounter on EVENT_DATE included ICD-10 code K85.8, Other acute pancreatitis<br>N=No; Y=Yes |
| K85_81 | Diagnoses recorded for the qualifying encounter on EVENT_DATE included ICD-10 code K85.81, Other acute pancreatitis with uninfected necrosis<br>N=No; Y=Yes |
| K85_9 | Diagnoses recorded for the qualifying encounter on EVENT_DATE included ICD-10 code K85.9, Acute pancreatitis, unspecified<br>N=No; Y=Yes |
| K85_90 | Diagnoses recorded for the qualifying encounter on EVENT_DATE included ICD-10 code K85.90, Acute pancreatitis without necrosis or infection, unspecified<br>N=No; Y=Yes |
| K85_91 | Diagnoses recorded for the qualifying encounter on EVENT_DATE included ICD-10 code K85.91, Acute pancreatitis with uninfected necrosis, unspecified<br>N=No; Y=Yes |
| K85_92 | Diagnoses recorded for the qualifying encounter on EVENT_DATE included ICD-10 code K85.92, Acute pancreatitis with infected necrosis, unspecified<br>N=No; Y=Yes |

| | |
|---|---|
| LIP14DT | Date maximum lipase result in the period +/-14 days around EVENT_DATE (a 29-day period) around event date occurred (e.g. "05mar2018") |
| FEMALE | Patient sex is female from administrative healthcare data.<br>0=No (if GENDER=M); 1=Yes (if GENDER=F) |
| ENCTYPE | Initial encounter type with AP code<br>1=IP; 2=ED; 3=AV |
| PRINCIPAL | For events where GROUP = IP, indicates whether the study-qualifying AP diagnosis on EVENT_DATE was recorded as the prinicpal diagnosis<br>0=No (if GROUP=IP and study-qualifying AP diagnosis is not principal diagnosis , or GROUP=ED or AV);<br>1=Yes (if GROUP=IP and study-qualifying AP diagnosis is principal diagnosis) |
| UNSPEC | Binary flag indicating whether the study-qualifying AP diagnosis on EVENT_DATE was coded as "unspecified" AP (i.e. any of the ICD-10 codes K85_8, K85_81, K85_9, K85_90, K85_91, K85_92)<br>0=No; 1=Yes |
| BILIARY | Binary flag indicating whether the study-qualifying AP diagnosis on EVENT_DATE was coded as "biliary" AP (i.e. any of the ICD-10 codes K85_1, K85_10)<br>0=No; 1=Yes |
| ETOH | Binary flag indicating whether the study-qualifying AP diagnosis on EVENT_DATE was coded as "alcohol induced" AP (i.e. any of the ICD-10 codes K85_2, K85_20)<br>0=No; 1=Yes |
| DRUG | Binary flag indicating whether the study-qualifying AP diagnosis on EVENT_DATE was coded as "drug induced" AP (i.e. ICD-10 code K85_3)<br>0=No; 1=Yes |
| NECROSIS_OUTCOME | Binary flag indicating whether the study-qualifying AP diagnosis on EVENT_DATE was coded as AP with uninfected necrosis (i.e. any of the ICD-10 codes K85_81, K85_91)<br>0=No; 1=Yes |
| INFNECROSIS | Binary flag indicating whether the study-qualifying AP diagnosis on EVENT_DATE was coded as AP with infected necrosis (i.e. ICD-10 code K85_92)<br>0=No; 1=Yes |
| LIP14ULN | Lipase Upper Limit of Normal in the period +/-14 days around EVENT_DATE (a 29-day period)<br>Note: if missing then no lab value available |
| LIP14X3 | Is Lipase in the period +/-14 days around EVENT_DATE (a 29-day period) 3 times or more than the Upper Limit of Normal?<br>0=No; 1=Yes<br>Note: if missing then no lab value available |

| | |
|---|---|
| LIP14X10 | Is Lipase in the period +/-14 days around EVENT_DATE (a 29-day period) 10 times or more than the Upper Limit of Normal?<br>0=No; 1=Yes<br>Note: if missing then no lab value available |
| ABDIMG | Abdominal imaging<br>0=No; 1=Yes |
| GROUP1 | Is sampling group IP at AP diagnosis? Also referred to as Group 1.<br>0=No; 1=Yes |
| GROUP2 | Is sampling group ED at AP diagnosis? Also referred to as Group 2.<br>0=No; 1=Yes |
| GROUP3 | Is sampling group OP/AV at AP diagnosis? Also referred to as Group 3.<br>0=No; 1=Yes |
| UPCLASS_EDIP | Is care setting up-classified to ED/IP?<br>0=No; 1=Yes<br>A binary flag indicating when the care setting of a study-qualifying encounter initially occurred in a lower-intensity care setting that was subsequently followed (within 7 days) by another study-qualifying encounter in a high-intensity setting. |
| ENRLYEARS | # years of continuous enrollment in Group Practice HMO prior to EVENTDTE_START<br>We define continuous enrollment as a period of enrollment ignoring gaps of up to 2 months (63 days) between 2 consecutive enrolled periods (because most such gaps are artifacts of administrative processes and do not represent actual disenrollment). Each patient will have a non-missing value and the minimum value will be 1. |
| AGE_AT_EVENT_IN_YRS | Age at EVENTDTE_START in years (integer) |
| RACE_UNK | Is race unknown?<br>0=No; 1=Yes |
| RACE_AIAN | Is race known to be American Indian or Alaska Native?<br>0=No; 1=Yes |
| RACE_ASIAN | Is race known to be Asian?<br>0=No; 1=Yes |
| RACE_AA | Is race known to be black or African-American?<br>0=No; 1=Yes |
| RACE_PI | Is race known to be Native Hawaiian or Other Pacific Islander?<br>0=No; 1=Yes |
| RACE_WHITE | Is race known to be white?<br>0=No; 1=Yes |

| | |
|---|---|
| HISPANIC | Hispanic ethnicity<br>N=No; U=Unknown; Y=Yes |
| SMOKE_CURR_SELF_365 | Patient is a current smoker per self-report during the past year<br>0=No; 1=Yes |
| SMOKE_FORMER_SELF_365 | Patient is a former smoker per self-report during the past year<br>0=No; 1=Yes |
| SMOKE_DX_PX_365 | Diagnosis or procedure code from any setting indicating current smoker status in the past year (i.e. during the 364-day period prior to or on EVENTDTE_START (365 days total)<br>0=No, 1=Yes |
| ALC_SD | Same day alcohol disorder<br>0=No; 1=Yes<br>Any alcohol disorder per HEDIS definition at encounter, the day before, or the day after |
| ALC_CS | Contemporaneous alcohol disorder<br>0=No; 1=Yes<br>Any alcohol disorder per HEDIS definition from 2 to 7 days before through 2 to 7 days after the encounter |
| ALC_PY | Past year alcohol disorder<br>0=No; 1=Yes<br>Any alcohol disorder per HEDIS definition from 8 to 365 days before the encounter |
| HYPERTRIG_SD | Same day familial hypertriglyceridemia<br>0=No; 1=Yes<br>Any significant hypertriglyceridemia at encounter, the day before, or the day after |
| HYPERTRIG_CS | Contemporaneous familial hypertriglyceridemia<br>0=No; 1=Yes<br>Any significant hypertriglyceridemia from 2 to 7 days before through 2 to 7 days after the encounter |
| HYPERTRIG_PY | Past year familial hypertriglyceridemia<br>0=No; 1=Yes<br>Any significant hypertriglyceridemia from 8 to 365 days before the encounter |
| ERCP | ERCP<br>0=No; 1=Yes<br>Any Endoscopic Retrograde Cholangiopancreatography within 14 days prior to EVENTDTE_START.. |
| ABDIMG_CT_14 | Abdominal imaging - CT scan from 14 days before through 14 days after EVENTDTE_START.<br>0=No; 1=Yes. |

| | |
|---|---|
| ABDIMG_MR_14 | Abdominal imaging - MRI from 14 days before through 14 days after EVENTDTE_START<br>0=No; 1=Yes |
| APLAB_MAX_ULN_14BEF_14AFT | Maximum lipase or amylase lab (normalized to the upper limit of normal) for all labs +/-14 days from EVENTDTE_START. |
| APLAB_MAX_GT3_14BEF_14AFT | This binary flag indicates whether the patient has any lipase (or amylase) lab(s) during the period +/-14 days from EVENTDTE_START that is greater than or equal to 3 times the upper limit of normal for that lab measurement.<br>0=No; 1=Yes |
| CHR_PANCR_SD | Same day chronic pancreatitis dx<br>0=No; 1=Yes<br>Any chronic pancreatitis dx at encounter, the day before, or the day after |
| CHR_PANCR_CS | Contemporaneous chronic pancreatitis dx<br>0=No; 1=Yes<br>Any chronic pancreatitis dx from 2 to 7 days before through 2 to 7 days after the encounter |
| CHR_PANCR_PY | Past year chronic pancreatitis dx<br>0=No; 1=Yes<br>Any chronic pancreatitis dx from 8 to 365 days before the encounter |
| PANC_CNCR_SD | Same day pancreatic cancer dx<br>0=No; 1=Yes<br>Any pancreatic cancer dx at encounter, the day before, or the day after |
| PANC_CNCR_CS | Contemporaneous pancreatic cancer dx<br>0=No; 1=Yes<br>Any pancreatic cancer dx from 2 to 7 days before through 2 to 7 days after the encounter |
| PANC_CNCR_PY | Past year pancreatic cancer dx<br>0=No; 1=Yes<br>Any pancreatic cancer dx from 8 to 365 days before the encounter |
| PEPTIC_ULCER_SD | Same day peptic ulcer disease dx<br>0=No; 1=Yes<br>Any peptic ulcer disease dx at encounter, the day before, or the day after |
| PEPTIC_ULCER_CS | Contemporaneous peptic ulcer disease dx<br>0=No; 1=Yes<br>Any peptic ulcer disease dx from 2 to 7 days before through 2 to 7 days after the encounter |

| | |
|---|---|
| PEPTIC_ULCER_PY | Past year peptic ulcer disease dx<br>0=No; 1=Yes<br>Any peptic ulcer disease dx from 8 to 365 days before the encounter |
| GASTRITIS_SD | Same day gastritis and duodenitis dx<br>0=No; 1=Yes<br>Any gastritis and duodenitis dx at encounter, the day before, or the day after |
| GASTRITIS_CS | Contemporaneous gastritis and duodenitis dx<br>0=No; 1=Yes<br>Any gastritis and duodenitis dx from 2 to 7 days before through 2 to 7 days after the encounter |
| GASTRITIS_PY | Past year gastritis and duodenitis dx<br>0=No; 1=Yes<br>Any gastritis and duodenitis dx from 8 to 365 days before the encounter |
| GERD_SD | Same day GERD dx<br>0=No; 1=Yes<br>Any GERD dx (Gastroesophageal reflux disease) at encounter, the day before, or the day after |
| GERD_CS | Contemporaneous GERD dx<br>0=No; 1=Yes<br>Any GERD dx (Gastroesophageal reflux disease) from 2 to 7 days before through 2 to 7 days after the encounter |
| GERD_PY | Past year GERD dx<br>0=No; 1=Yes<br>Any GERD dx (Gastroesophageal reflux disease) from 8 to 365 days before the encounter |
| INTEST_OBS_SD | Same day intestinal obstruction dx<br>0=No; 1=Yes<br>Any intestinal obstruction dx at encounter, the day before, or the day after |
| INTEST_OBS_CS | Contemporaneous intestinal obstruction dx<br>0=No; 1=Yes<br>Any intestinal obstruction dx from 2 to 7 days before through 2 to 7 days after the encounter |
| INTEST_OBS_PY | Past year intestinal obstruction dx<br>0=No; 1=Yes<br>Any intestinal obstruction dx from 8 to 365 days before the encounter |

| | |
|---|---|
| ILEUS_SD | Same day ileus dx<br>0=No; 1=Yes<br>Any Ileus dx at encounter, the day before, or the day after |
| ILEUS_CS | Contemporaneous ileus dx<br>0=No; 1=Yes<br>Any Ileus dx from 2 to 7 days before through 2 to 7 days after the encounter |
| ILEUS_PY | Past year ileus dx<br>0=No; 1=Yes<br>Any Ileus dx from 8 to 365 days before the encounter |
| CONSTIPATION_SD | Same day constipation dx<br>0=No; 1=Yes<br>Any constipation dx at encounter, the day before, or the day after |
| CONSTIPATION_CS | Contemporaneous constipation dx<br>0=No; 1=Yes<br>Any constipation dx from 2 to 7 days before through 2 to 7 days after the encounter |
| CONSTIPATION_PY | Past year constipation dx<br>0=No; 1=Yes<br>Any constipation dx from 8 to 365 days before the encounter |
| MESENT_ISCH_SD | Same day mesenteric ischemia dx<br>0=No; 1=Yes<br>Any mesenteric ischemia dx at encounter, the day before, or the day after |
| MESENT_ISCH_CS | Contemporaneous mesenteric ischemia dx<br>0=No; 1=Yes<br>Any mesenteric ischemia dx from 2 to 7 days before through 2 to 7 days after the encounter |
| MESENT_ISCH_PY | Past year mesenteric ischemia dx<br>0=No; 1=Yes<br>Any mesenteric ischemia dx from 8 to 365 days before the encounter |
| DIVERTICUL_SD | Same day diverticulitis disease dx<br>0=No; 1=Yes<br>Any diverticulitis disease dx at encounter, the day before, or the day after |
| DIVERTICUL_CS | Contemporaneous diverticulitis disease dx<br>0=No; 1=Yes<br>Any diverticulitis disease dx from 2 to 7 days before through 2 to 7 days after the encounter |

| | |
|---|---|
| DIVERTICUL_PY | Past year diverticulitis disease dx<br>0=No; 1=Yes<br>Any diverticulitis disease dx from 8 to 365 days before the encounter |
| APPENDIC_SD | Same day appendicitis dx<br>0=No; 1=Yes<br>Any appendicitis dx at encounter, the day before, or the day after |
| APPENDIC_CS | Contemporaneous appendicitis dx<br>0=No; 1=Yes<br>Any appendicitis dx from 2 to 7 days before through 2 to 7 days after the encounter |
| APPENDIC_PY | Past year appendicitis dx<br>0=No; 1=Yes<br>Any appendicitis dx from 8 to 365 days before the encounter |
| HEPATITIS_SD | Same day hepatitis dx<br>0=No; 1=Yes<br>Any hepatitis dx at encounter, the day before, or the day after |
| HEPATITIS_CS | Contemporaneous hepatitis dx<br>0=No; 1=Yes<br>Any hepatitis dx from 2 to 7 days before through 2 to 7 days after the encounter |
| HEPATITIS_PY | Past year hepatitis dx<br>0=No; 1=Yes<br>Any hepatitis dx from 8 to 365 days before the encounter |
| INFLUENZA_SD | Same day influenza dx<br>0=No; 1=Yes<br>Any influenza dx at encounter, the day before, or the day after |
| INFLUENZA_CS | Contemporaneous influenza dx<br>0=No; 1=Yes<br>Any influenza dx from 2 to 7 days before through 2 to 7 days after the encounter |
| INFLUENZA_PY | Past year influenza dx<br>0=No; 1=Yes<br>Any influenza dx from 8 to 365 days before the encounter |
| FOOD_POIS_SD | Same day food poisoning dx<br>0=No; 1=Yes<br>Any food poisoning dx at encounter, the day before, or the day after |
| FOOD_POIS_CS | Contemporaneous food poisoning dx<br>0=No; 1=Yes<br>Any food poisoning dx from 2 to 7 days before through 2 to 7 days after the encounter |

| | |
|---|---|
| FOOD_POIS_PY | Past year food poisoning dx<br>0=No; 1=Yes<br>Any food poisoning dx from 8 to 365 days before the encounter |
| ASCITES_SD | Same day ascites dx<br>0=No; 1=Yes<br>Any ascites dx at encounter, the day before, or the day after |
| ASCITES_CS | Contemporaneous ascites dx<br>0=No; 1=Yes<br>Any ascites dx from 2 to 7 days before through 2 to 7 days after the encounter |
| ASCITES_PY | Past year ascites dx<br>0=No; 1=Yes<br>Any ascites dx from 8 to 365 days before the encounter |
| NEPHROLITH_SD | Same day nephrolithiasis dx<br>0=No; 1=Yes<br>Any nephrolithiasis dx at encounter, the day before, or the day after |
| NEPHROLITH_CS | Contemporaneous nephrolithiasis dx<br>0=No; 1=Yes<br>Any nephrolithiasis dx from 2 to 7 days before through 2 to 7 days after the encounter |
| NEPHROLITH_PY | Past year nephrolithiasis dx<br>0=No; 1=Yes<br>Any nephrolithiasis dx from 8 to 365 days before the encounter |
| DKA_SD | Same day DKA dx<br>0=No; 1=Yes<br>Any DKA dx (Diabetic ketoacidosis) at encounter, the day before, or the day after |
| DKA_CS | Contemporaneous DKA dx<br>0=No; 1=Yes<br>Any DKA dx (Diabetic ketoacidosis) from 2 to 7 days before through 2 to 7 days after the encounter |
| DKA_PY | Past year DKA dx<br>0=No; 1=Yes<br>Any DKA dx (Diabetic ketoacidosis) from 8 to 365 days before the encounter |
| MYOCARD_ISCH_SD | Same day myocardial ischemia dx<br>0=No; 1=Yes<br>Any myocardial ischemia dx at encounter, the day before, or the day after |

| | |
|---|---|
| MYOCARD_ISCH_CS | Contemporaneous myocardial ischemia dx<br>0=No; 1=Yes<br>Any myocardial ischemia dx from 2 to 7 days before through 2 to 7 days after the encounter |
| MYOCARD_ISCH_PY | Past year myocardial ischemia dx<br>0=No; 1=Yes<br>Any myocardial ischemia dx from 8 to 365 days before the encounter |
| GALL_BIL_SD | Same day gallbladder and biliary disease dx<br>0=No; 1=Yes<br>Any gallbladder and biliary disease dx at encounter, the day before, or the day after |
| GALL_BIL_CS | Contemporaneous gallbladder and biliary disease dx<br>0=No; 1=Yes<br>Any gallbladder and biliary disease dx from 2 to 7 days before through 2 to 7 days after the encounter |
| GALL_BIL_PY | Past year gallbladder and biliary disease dx<br>0=No; 1=Yes<br>Any gallbladder and biliary disease dx from 8 to 365 days before the encounter |
| GB_CANCER_SD | Same day cancer of gallbladder and biliary tract dx<br>0=No; 1=Yes<br>Any dx of cancer of gallbladder and biliary tract at encounter, the day before, or the day after |
| GB_CANCER_CS | Contemporaneous cancer of gallbladder and biliary tract dx<br>0=No; 1=Yes<br>Any dx of cancer of gallbladder and biliary tract from 2 to 7 days before through 2 to 7 days after the encounter |
| GB_CANCER_PY | Past year cancer of gallbladder and biliary tract dx<br>0=No; 1=Yes<br>Any dx of cancer of gallbladder and biliary tract from 8 to 365 days before the encounter |
| IBD_SD | Same day inflammatory bowel disease dx<br>0=No; 1=Yes<br>Any inflammatory bowel disease dx at encounter, the day before, or the day after |
| IBD_CS | Contemporaneous inflammatory bowel disease dx<br>0=No; 1=Yes<br>Any inflammatory bowel disease dx from 2 to 7 days before through 2 to 7 days after the encounter |

| | |
|---|---|
| IBD_PY | Past year inflammatory bowel disease dx<br>0=No; 1=Yes<br>Any inflammatory bowel disease dx from 8 to 365 days before the encounter |
| GASTRO_SD | Same day infectious gastroenteritis and colitis dx<br>0=No; 1=Yes<br>Any infectious gastroenteritis and colitis dx at encounter, the day before, or the day after |
| GASTRO_CS | Contemporaneous infectious gastroenteritis and colitis dx<br>0=No; 1=Yes<br>Any infectious gastroenteritis and colitis dx from 2 to 7 days before through 2 to 7 days after the encounter |
| GASTRO_PY | Past year infectious gastroenteritis and colitis dx<br>0=No; 1=Yes<br>Any infectious gastroenteritis and colitis dx from 8 to 365 days before the encounter |
| ESOPHAGITIS_SD | Same day esophagitis dx<br>0=No; 1=Yes<br>Any esophagitis dx at encounter, the day before, or the day after |
| ESOPHAGITIS_CS | Contemporaneous esophagitis dx<br>0=No; 1=Yes<br>Any esophagitis dx from 2 to 7 days before through 2 to 7 days after the encounter |
| ESOPHAGITIS_PY | Past year esophagitis dx<br>0=No; 1=Yes<br>Any esophagitis dx from 8 to 365 days before the encounter |

### Appendix B: Manually Curated NLP-Derived Covariates

In collaboration between clinicians and informaticists, we manually engineered the following candidate NLP covariates.

| Covariate Name | Description |
|---|---|
| patient_id | Unique arbitrary identifier of patient/potential event |
| pain | Any pain mention |
| pain_all | Any pain mention (all mentions counted) (all notes) |
| pain_is_r | Any pain mention (number of encounters with mentions) (radiology/imaging only) |
| pain_all_is_r | Any pain mention (all mentions counted) (radiology/imaging only) |
| competing_dx | Any competing diagnoses mention |
| competing_dx_all | Any competing diagnoses mention (all mentions counted) (all notes) |
| competing_dx_is_r | Any competing diagnoses mention (number of encounters with mentions) (radiology/imaging only) |
| competing_dx_all_is_r | Any competing diagnoses mention (all mentions counted) (radiology/imaging only) |
| pancreatitis | Any pancreatitis mention |
| pancreatitis_all | Any pancreatitis mention (all mentions counted) (all notes) |
| pancreatitis_is_r | Any pancreatitis mention (number of encounters with mentions) (radiology/imaging only) |
| pancreatitis_all_is_r | Any pancreatitis mention (all mentions counted) (radiology/imaging only) |
| nausea | Any nausea mention |
| nausea_all | Any nausea mention (all mentions counted) (all notes) |
| nausea_is_r | Any nausea mention (number of encounters with mentions) (radiology/imaging only) |
| nausea_all_is_r | Any nausea mention (all mentions counted) (radiology/imaging only) |
| necrosis | Any necrosis mention |
| necrosis_all | Any necrosis mention (all mentions counted) (all notes) |
| necrosis_is_r | Any necrosis mention (number of encounters with mentions) (radiology/imaging only) |
| necrosis_all_is_r | Any necrosis mention (all mentions counted) (radiology/imaging only) |
| fluid | Any fluid mention |
| fluid_all | Any fluid mention (all mentions counted) (all notes) |
| fluid_is_r | Any fluid mention (number of encounters with mentions) (radiology/imaging only) |
| fluid_all_is_r | Any fluid mention (all mentions counted) (radiology/imaging only) |
| pseudocyst | Any pseudocyst mention |
| pseudocyst_all | Any pseudocyst mention (all mentions counted) (all notes) |

| | |
|---|---|
| pseudocyst_is_r | Any pseudocyst mention (number of encounters with mentions) (radiology/imaging only) |
| pseudocyst_all_is_r | Any pseudocyst mention (all mentions counted) (radiology/imaging only) |
| pain_EPIGASTRIC | Epigastric pain |
| pain_EPIGASTRIC_all | Epigastric pain (all mentions counted) (all notes) |
| pain_EPIGASTRIC_is_r | Epigastric pain (number of encounters with mentions) (radiology/imaging only) |
| pain_EPIGASTRIC_all_is_r | Epigastric pain (all mentions counted) (radiology/imaging only) |
| pain_CHEST | Chest pain |
| pain_CHEST_all | Chest pain (all mentions counted) (all notes) |
| pain_CHEST_is_r | Chest pain (number of encounters with mentions) (radiology/imaging only) |
| pain_CHEST_all_is_r | Chest pain (all mentions counted) (radiology/imaging only) |
| competing_dx_NEGATIVE | Presence of negative (competing diagnosis) |
| competing_dx_NEGATIVE_all | Presence of negative (competing diagnosis) (all mentions counted) (all notes) |
| competing_dx_NEGATIVE_is_r | Presence of negative (competing diagnosis) (number of encounters with mentions) (radiology/imaging only) |
| competing_dx_NEGATIVE_all_is_r | Presence of negative (competing diagnosis) (all mentions counted) (radiology/imaging only) |
| competing_dx_GERD | Presence of gerd (competing diagnosis) |
| competing_dx_GERD_all | Presence of gerd (competing diagnosis) (all mentions counted) (all notes) |
| competing_dx_GERD_is_r | Presence of gerd (competing diagnosis) (number of encounters with mentions) (radiology/imaging only) |
| competing_dx_GERD_all_is_r | Presence of gerd (competing diagnosis) (all mentions counted) (radiology/imaging only) |
| competing_dx_IBD | Presence of ibd (competing diagnosis) |
| competing_dx_IBD_all | Presence of ibd (competing diagnosis) (all mentions counted) (all notes) |
| competing_dx_IBD_is_r | Presence of ibd (competing diagnosis) (number of encounters with mentions) (radiology/imaging only) |
| competing_dx_IBD_all_is_r | Presence of ibd (competing diagnosis) (all mentions counted) (radiology/imaging only) |
| pancreatitis_ACUTE | Acute pancreatitis |
| pancreatitis_ACUTE_all | Acute pancreatitis (all mentions counted) (all notes) |
| pancreatitis_ACUTE_is_r | Acute pancreatitis (number of encounters with mentions) (radiology/imaging only) |
| pancreatitis_ACUTE_all_is_r | Acute pancreatitis (all mentions counted) (radiology/imaging only) |
| pancreatitis_CONSISTENT_WITH | Radiology conclusion-like language (e.g., consistent with) |
| pancreatitis_CONSISTENT_WITH_all | Radiology conclusion-like language (e.g., consistent with) (all mentions counted) (all notes) |
| pancreatitis_CONSISTENT_WITH_is_r | Radiology conclusion-like language (e.g., consistent with) (number of encounters with mentions) (radiology/imaging only) |

| | |
|---|---|
| pancreatitis_CONSISTENT_WITH_all_is_r | Radiology conclusion-like language (e.g., consistent with) (all mentions counted) (radiology/imaging only) |
| pain_RADIATING_TO_BACK | Pain described as radiating to the back |
| pain_RADIATING_TO_BACK_all | Pain described as radiating to the back (all mentions counted) (all notes) |
| pain_RADIATING_TO_BACK_is_r | Pain described as radiating to the back (number of encounters with mentions) (radiology/imaging only) |
| pain_RADIATING_TO_BACK_all_is_r | Pain described as radiating to the back (all mentions counted) (radiology/imaging only) |
| nausea_VOMITING | Vomiting |
| nausea_VOMITING_all | Vomiting (all mentions counted) (all notes) |
| nausea_VOMITING_is_r | Vomiting (number of encounters with mentions) (radiology/imaging only) |
| nausea_VOMITING_all_is_r | Vomiting (all mentions counted) (radiology/imaging only) |
| nausea_NAUSEA | Nausea |
| nausea_NAUSEA_all | Nausea (all mentions counted) (all notes) |
| nausea_NAUSEA_is_r | Nausea (number of encounters with mentions) (radiology/imaging only) |
| nausea_NAUSEA_all_is_r | Nausea (all mentions counted) (radiology/imaging only) |
| pain_ABD_PAIN | Abdominal pain |
| pain_ABD_PAIN_all | Abdominal pain (all mentions counted) (all notes) |
| pain_ABD_PAIN_is_r | Abdominal pain (number of encounters with mentions) (radiology/imaging only) |
| pain_ABD_PAIN_all_is_r | Abdominal pain (all mentions counted) (radiology/imaging only) |
| competing_dx_INFLUENZA | Presence of influenza (competing diagnosis) |
| competing_dx_INFLUENZA_all | Presence of influenza (competing diagnosis) (all mentions counted) (all notes) |
| competing_dx_INFLUENZA_is_r | Presence of influenza (competing diagnosis) (number of encounters with mentions) (radiology/imaging only) |
| competing_dx_INFLUENZA_all_is_r | Presence of influenza (competing diagnosis) (all mentions counted) (radiology/imaging only) |
| pancreatitis_POSITIVE | Generic pancreatitis |
| pancreatitis_POSITIVE_all | Generic pancreatitis (all mentions counted) (all notes) |
| pancreatitis_POSITIVE_is_r | Generic pancreatitis (number of encounters with mentions) (radiology/imaging only) |
| pancreatitis_POSITIVE_all_is_r | Generic pancreatitis (all mentions counted) (radiology/imaging only) |
| competing_dx_ASCITES | Presence of ascites (competing diagnosis) |
| competing_dx_ASCITES_all | Presence of ascites (competing diagnosis) (all mentions counted) (all notes) |
| competing_dx_ASCITES_is_r | Presence of ascites (competing diagnosis) (number of encounters with mentions) (radiology/imaging only) |
| competing_dx_ASCITES_all_is_r | Presence of ascites (competing diagnosis) (all mentions counted) (radiology/imaging only) |
| competing_dx_HEPATITIS | Presence of hepatitis (competing diagnosis) |

| | |
|---|---|
| competing_dx_HEPATITIS_all | Presence of hepatitis (competing diagnosis) (all mentions counted) (all notes) |
| competing_dx_HEPATITIS_is_r | Presence of hepatitis (competing diagnosis) (number of encounters with mentions) (radiology/imaging only) |
| competing_dx_HEPATITIS_all_is_r | Presence of hepatitis (competing diagnosis) (all mentions counted) (radiology/imaging only) |
| pain_ACUTE | Acute pain |
| pain_ACUTE_all | Acute pain (all mentions counted) (all notes) |
| pain_ACUTE_is_r | Acute pain (number of encounters with mentions) (radiology/imaging only) |
| pain_ACUTE_all_is_r | Acute pain (all mentions counted) (radiology/imaging only) |
| competing_dx_INFECTIOUS_GE | Presence of infectious gastroenteritis (competing diagnosis) |
| competing_dx_INFECTIOUS_GE_all | Presence of infectious gastroenteritis (competing diagnosis) (all mentions counted) (all notes) |
| competing_dx_INFECTIOUS_GE_is_r | Presence of infectious gastroenteritis (competing diagnosis) (number of encounters with mentions) (radiology/imaging only) |
| competing_dx_INFECTIOUS_GE_all_is_r | Presence of infectious gastroenteritis (competing diagnosis) (all mentions counted) (radiology/imaging only) |
| competing_dx_PEPTIC_ULCER | Presence of peptic ulcer (competing diagnosis) |
| competing_dx_PEPTIC_ULCER_all | Presence of peptic ulcer (competing diagnosis) (all mentions counted) (all notes) |
| competing_dx_PEPTIC_ULCER_is_r | Presence of peptic ulcer (competing diagnosis) (number of encounters with mentions) (radiology/imaging only) |
| competing_dx_PEPTIC_ULCER_all_is_r | Presence of peptic ulcer (competing diagnosis) (all mentions counted) (radiology/imaging only) |
| pancreatitis_CHRONIC | Chronic pancreatitis |
| pancreatitis_CHRONIC_all | Chronic pancreatitis (all mentions counted) (all notes) |
| pancreatitis_CHRONIC_is_r | Chronic pancreatitis (number of encounters with mentions) (radiology/imaging only) |
| pancreatitis_CHRONIC_all_is_r | Chronic pancreatitis (all mentions counted) (radiology/imaging only) |
| competing_dx_BOWEL_MOVEMENTS | Presence of bowel_movements (competing diagnosis) |
| competing_dx_BOWEL_MOVEMENTS_all | Presence of bowel_movements (competing diagnosis) (all mentions counted) (all notes) |
| competing_dx_BOWEL_MOVEMENTS_is_r | Presence of bowel_movements (competing diagnosis) (number of encounters with mentions) (radiology/imaging only) |
| competing_dx_BOWEL_MOVEMENTS_all_is_r | Presence of bowel_movements (competing diagnosis) (all mentions counted) (radiology/imaging only) |
| competing_dx_BLOOD_IN_STOOL | Presence of blood in stool (competing diagnosis) |
| competing_dx_BLOOD_IN_STOOL_all | Presence of blood in stool (competing diagnosis) (all mentions counted) (all notes) |
| competing_dx_BLOOD_IN_STOOL_is_r | Presence of blood in stool (competing diagnosis) (number of encounters with mentions) (radiology/imaging only) |
| competing_dx_BLOOD_IN_STOOL_all_is_r | Presence of blood in stool (competing diagnosis) (all mentions counted) (radiology/imaging only) |

| | |
|---|---|
| pain_VERY_RECENT | Recent pain (in days) |
| pain_VERY_RECENT_all | Recent pain (in days) (all mentions counted) (all notes) |
| pain_VERY_RECENT_is_r | Recent pain (in days) (number of encounters with mentions) (radiology/imaging only) |
| pain_VERY_RECENT_all_is_r | Recent pain (in days) (all mentions counted) (radiology/imaging only) |
| competing_dx_STROKE | Presence of stroke (competing diagnosis) |
| competing_dx_STROKE_all | Presence of stroke (competing diagnosis) (all mentions counted) (all notes) |
| competing_dx_STROKE_is_r | Presence of stroke (competing diagnosis) (number of encounters with mentions) (radiology/imaging only) |
| competing_dx_STROKE_all_is_r | Presence of stroke (competing diagnosis) (all mentions counted) (radiology/imaging only) |
| competing_dx_COLANGITIS | Presence of colangitis (competing diagnosis) |
| competing_dx_COLANGITIS_all | Presence of colangitis (competing diagnosis) (all mentions counted) (all notes) |
| competing_dx_COLANGITIS_is_r | Presence of colangitis (competing diagnosis) (number of encounters with mentions) (radiology/imaging only) |
| competing_dx_COLANGITIS_all_is_r | Presence of colangitis (competing diagnosis) (all mentions counted) (radiology/imaging only) |
| competing_dx_CONSTIPATION | Presence of constipation (competing diagnosis) |
| competing_dx_CONSTIPATION_all | Presence of constipation (competing diagnosis) (all mentions counted) (all notes) |
| competing_dx_CONSTIPATION_is_r | Presence of constipation (competing diagnosis) (number of encounters with mentions) (radiology/imaging only) |
| competing_dx_CONSTIPATION_all_is_r | Presence of constipation (competing diagnosis) (all mentions counted) (radiology/imaging only) |
| necrosis_ACUTE_COLLECTION | Acute necrotic collection |
| necrosis_ACUTE_COLLECTION_all | Acute necrotic collection (all mentions counted) (all notes) |
| necrosis_ACUTE_COLLECTION_is_r | Acute necrotic collection (number of encounters with mentions) (radiology/imaging only) |
| necrosis_ACUTE_COLLECTION_all_is_r | Acute necrotic collection (all mentions counted) (radiology/imaging only) |
| pain_RECENT | Recent pain (in weeks) |
| pain_RECENT_all | Recent pain (in weeks) (all mentions counted) (all notes) |
| pain_RECENT_is_r | Recent pain (in weeks) (number of encounters with mentions) (radiology/imaging only) |
| pain_RECENT_all_is_r | Recent pain (in weeks) (all mentions counted) (radiology/imaging only) |
| pain_CHRONIC | Chronic pain |
| pain_CHRONIC_all | Chronic pain (all mentions counted) (all notes) |
| pain_CHRONIC_is_r | Chronic pain (number of encounters with mentions) (radiology/imaging only) |
| pain_CHRONIC_all_is_r | Chronic pain (all mentions counted) (radiology/imaging only) |
| competing_dx_NEPHROLITHIASIS | Presence of nephrolithiasis (competing diagnosis) |

| | |
|---|---|
| competing_dx_NEPHROLITHIASIS_all | Presence of nephrolithiasis (competing diagnosis) (all mentions counted) (all notes) |
| competing_dx_NEPHROLITHIASIS_is_r | Presence of nephrolithiasis (competing diagnosis) (number of encounters with mentions) (radiology/imaging only) |
| competing_dx_NEPHROLITHIASIS_all_is_r | Presence of nephrolithiasis (competing diagnosis) (all mentions counted) (radiology/imaging only) |
| pancreatitis_INFLAMMATION | Inflammation |
| pancreatitis_INFLAMMATION_all | Inflammation (all mentions counted) (all notes) |
| pancreatitis_INFLAMMATION_is_r | Inflammation (number of encounters with mentions) (radiology/imaging only) |
| pancreatitis_INFLAMMATION_all_is_r | Inflammation (all mentions counted) (radiology/imaging only) |
| pancreatitis_PERI_INFLAMMATION | Peri-inflammation |
| pancreatitis_PERI_INFLAMMATION_all | Peri-inflammation (all mentions counted) (all notes) |
| pancreatitis_PERI_INFLAMMATION_is_r | Peri-inflammation (number of encounters with mentions) (radiology/imaging only) |
| pancreatitis_PERI_INFLAMMATION_all_is_r | Peri-inflammation (all mentions counted) (radiology/imaging only) |
| pain_SUDDEN_ONSET | Sudden onset pain |
| pain_SUDDEN_ONSET_all | Sudden onset pain (all mentions counted) (all notes) |
| pain_SUDDEN_ONSET_is_r | Sudden onset pain (number of encounters with mentions) (radiology/imaging only) |
| pain_SUDDEN_ONSET_all_is_r | Sudden onset pain (all mentions counted) (radiology/imaging only) |
| competing_dx_DIVERTICULOSIS | Presence of diverticulosis (competing diagnosis) |
| competing_dx_DIVERTICULOSIS_all | Presence of diverticulosis (competing diagnosis) (all mentions counted) (all notes) |
| competing_dx_DIVERTICULOSIS_is_r | Presence of diverticulosis (competing diagnosis) (number of encounters with mentions) (radiology/imaging only) |
| competing_dx_DIVERTICULOSIS_all_is_r | Presence of diverticulosis (competing diagnosis) (all mentions counted) (radiology/imaging only) |
| fluid_PANCREATIC | Pancreatic fluid |
| fluid_PANCREATIC_all | Pancreatic fluid (all mentions counted) (all notes) |
| fluid_PANCREATIC_is_r | Pancreatic fluid (number of encounters with mentions) (radiology/imaging only) |
| fluid_PANCREATIC_all_is_r | Pancreatic fluid (all mentions counted) (radiology/imaging only) |
| pain_UNKNOWN_DURATION | Likely unrecognized time format for recency |
| pain_UNKNOWN_DURATION_all | Likely unrecognized time format for recency (all mentions counted) (all notes) |
| pain_UNKNOWN_DURATION_is_r | Likely unrecognized time format for recency (number of encounters with mentions) (radiology/imaging only) |
| pain_UNKNOWN_DURATION_all_is_r | Likely unrecognized time format for recency (all mentions counted) (radiology/imaging only) |
| competing_dx_CHRONIC_PAIN | Presence of chronic pain (competing diagnosis) |

| | |
|---|---|
| competing_dx_CHRONIC_PAIN_all | Presence of chronic pain (competing diagnosis) (all mentions counted) (all notes) |
| competing_dx_CHRONIC_PAIN_is_r | Presence of chronic pain (competing diagnosis) (number of encounters with mentions) (radiology/imaging only) |
| competing_dx_CHRONIC_PAIN_all_is_r | Presence of chronic pain (competing diagnosis) (all mentions counted) (radiology/imaging only) |
| pseudocyst_PANCREATIC | Pancreatic pseudocyst |
| pseudocyst_PANCREATIC_all | Pancreatic pseudocyst (all mentions counted) (all notes) |
| pseudocyst_PANCREATIC_is_r | Pancreatic pseudocyst (number of encounters with mentions) (radiology/imaging only) |
| pseudocyst_PANCREATIC_all_is_r | Pancreatic pseudocyst (all mentions counted) (radiology/imaging only) |
| pseudocyst_POSITIVE | Generic pseudocyst |
| pseudocyst_POSITIVE_all | Generic pseudocyst (all mentions counted) (all notes) |
| pseudocyst_POSITIVE_is_r | Generic pseudocyst (number of encounters with mentions) (radiology/imaging only) |
| pseudocyst_POSITIVE_all_is_r | Generic pseudocyst (all mentions counted) (radiology/imaging only) |
| competing_dx_GALL_BLADDER_DISEASE | Presence of gall bladder disease (competing diagnosis) |
| competing_dx_GALL_BLADDER_DISEASE_all | Presence of gall bladder disease (competing diagnosis) (all mentions counted) (all notes) |
| competing_dx_GALL_BLADDER_DISEASE_is_r | Presence of gall bladder disease (competing diagnosis) (number of encounters with mentions) (radiology/imaging only) |
| competing_dx_GALL_BLADDER_DISEASE_all_is_r | Presence of gall bladder disease (competing diagnosis) (all mentions counted) (radiology/imaging only) |
| competing_dx_INTESTINAL_OBSTRUCTION | Presence of intestinal_obstruction (competing diagnosis) |
| competing_dx_INTESTINAL_OBSTRUCTION_all | Presence of intestinal_obstruction (competing diagnosis) (all mentions counted) (all notes) |
| competing_dx_INTESTINAL_OBSTRUCTION_is_r | Presence of intestinal_obstruction (competing diagnosis) (number of encounters with mentions) (radiology/imaging only) |
| competing_dx_INTESTINAL_OBSTRUCTION_all_is_r | Presence of intestinal_obstruction (competing diagnosis) (all mentions counted) (radiology/imaging only) |
| competing_dx_MESENTERIC_ISCHEMIA | Presence of mesenteric ischemia (competing diagnosis) |
| competing_dx_MESENTERIC_ISCHEMIA_all | Presence of mesenteric ischemia (competing diagnosis) (all mentions counted) (all notes) |
| competing_dx_MESENTERIC_ISCHEMIA_is_r | Presence of mesenteric ischemia (competing diagnosis) (number of encounters with mentions) (radiology/imaging only) |
| competing_dx_MESENTERIC_ISCHEMIA_all_is_r | Presence of mesenteric ischemia (competing diagnosis) (all mentions counted) (radiology/imaging only) |
| competing_dx_ILEUS | Presence of ileus (competing diagnosis) |
| competing_dx_ILEUS_all | Presence of ileus (competing diagnosis) (all mentions counted) (all notes) |

| | |
|---|---|
| competing_dx_ILEUS_is_r | Presence of ileus (competing diagnosis) (number of encounters with mentions) (radiology/imaging only) |
| competing_dx_ILEUS_all_is_r | Presence of ileus (competing diagnosis) (all mentions counted) (radiology/imaging only) |
| pain_WORSENING | Pain described as worsening |
| pain_WORSENING_all | Pain described as worsening (all mentions counted) (all notes) |
| pain_WORSENING_is_r | Pain described as worsening (number of encounters with mentions) (radiology/imaging only) |
| pain_WORSENING_all_is_r | Pain described as worsening (all mentions counted) (radiology/imaging only) |
| competing_dx_MYOCARDIAL_ISCHEMIA | Presence of myocardial ischemia (competing diagnosis) |
| competing_dx_MYOCARDIAL_ISCHEMIA_all | Presence of myocardial ischemia (competing diagnosis) (all mentions counted) (all notes) |
| competing_dx_MYOCARDIAL_ISCHEMIA_is_r | Presence of myocardial ischemia (competing diagnosis) (number of encounters with mentions) (radiology/imaging only) |
| competing_dx_MYOCARDIAL_ISCHEMIA_all_is_r | Presence of myocardial ischemia (competing diagnosis) (all mentions counted) (radiology/imaging only) |
| competing_dx_APPENDICITIS | Presence of appendicitis (competing diagnosis) |
| competing_dx_APPENDICITIS_all | Presence of appendicitis (competing diagnosis) (all mentions counted) (all notes) |
| competing_dx_APPENDICITIS_is_r | Presence of appendicitis (competing diagnosis) (number of encounters with mentions) (radiology/imaging only) |
| competing_dx_APPENDICITIS_all_is_r | Presence of appendicitis (competing diagnosis) (all mentions counted) (radiology/imaging only) |
| competing_dx_BLOOD_IN_VOMIT | Presence of blood in vomit (competing diagnosis) |
| competing_dx_BLOOD_IN_VOMIT_all | Presence of blood in vomit (competing diagnosis) (all mentions counted) (all notes) |
| competing_dx_BLOOD_IN_VOMIT_is_r | Presence of blood in vomit (competing diagnosis) (number of encounters with mentions) (radiology/imaging only) |
| competing_dx_BLOOD_IN_VOMIT_all_is_r | Presence of blood in vomit (competing diagnosis) (all mentions counted) (radiology/imaging only) |
| competing_dx_DKA | Presence of dka (competing diagnosis) |
| competing_dx_DKA_all | Presence of dka (competing diagnosis) (all mentions counted) (all notes) |
| competing_dx_DKA_is_r | Presence of dka (competing diagnosis) (number of encounters with mentions) (radiology/imaging only) |
| competing_dx_DKA_all_is_r | Presence of dka (competing diagnosis) (all mentions counted) (radiology/imaging only) |
| necrosis_POSITIVE | Generic mention of necrosis |
| necrosis_POSITIVE_all | Generic mention of necrosis (all mentions counted) (all notes) |
| necrosis_POSITIVE_is_r | Generic mention of necrosis (number of encounters with mentions) (radiology/imaging only) |
| necrosis_POSITIVE_all_is_r | Generic mention of necrosis (all mentions counted) (radiology/imaging only) |
| competing_dx_FOOD_POISONING | Presence of food poisoning (competing diagnosis) |

| | |
|---|---|
| competing_dx_FOOD_POISONING_all | Presence of food poisoning (competing diagnosis) (all mentions counted) (all notes) |
| competing_dx_FOOD_POISONING_is_r | Presence of food poisoning (competing diagnosis) (number of encounters with mentions) (radiology/imaging only) |
| competing_dx_FOOD_POISONING_all_is_r | Presence of food poisoning (competing diagnosis) (all mentions counted) (radiology/imaging only) |
| competing_dx_ACUTE_APPENDICITIS | Presence of acute appendicitis (competing diagnosis) |
| competing_dx_ACUTE_APPENDICITIS_all | Presence of acute appendicitis (competing diagnosis) (all mentions counted) (all notes) |
| competing_dx_ACUTE_APPENDICITIS_is_r | Presence of acute appendicitis (competing diagnosis) (number of encounters with mentions) (radiology/imaging only) |
| competing_dx_ACUTE_APPENDICITIS_all_is_r | Presence of acute appendicitis (competing diagnosis) (all mentions counted) (radiology/imaging only) |
| pain_LONG_AGO | Non-recent pain (in months) |
| pain_LONG_AGO_all | Non-recent pain (in months) (all mentions counted) (all notes) |
| pain_LONG_AGO_is_r | Non-recent pain (in months) (number of encounters with mentions) (radiology/imaging only) |
| pain_LONG_AGO_all_is_r | Non-recent pain (in months) (all mentions counted) (radiology/imaging only) |
| competing_dx_GASTRODUODENITIS | Presence of gastroduodenitis (competing diagnosis) |
| competing_dx_GASTRODUODENITIS_all | Presence of gastroduodenitis (competing diagnosis) (all mentions counted) (all notes) |
| competing_dx_GASTRODUODENITIS_is_r | Presence of gastroduodenitis (competing diagnosis) (number of encounters with mentions) (radiology/imaging only) |
| competing_dx_GASTRODUODENITIS_all_is_r | Presence of gastroduodenitis (competing diagnosis) (all mentions counted) (radiology/imaging only) |
| competing_dx_ESOPHAGITIS | Presence of esophagitis (competing diagnosis) |
| competing_dx_ESOPHAGITIS_all | Presence of esophagitis (competing diagnosis) (all mentions counted) (all notes) |
| competing_dx_ESOPHAGITIS_is_r | Presence of esophagitis (competing diagnosis) (number of encounters with mentions) (radiology/imaging only) |
| competing_dx_ESOPHAGITIS_all_is_r | Presence of esophagitis (competing diagnosis) (all mentions counted) (radiology/imaging only) |
| pancreatitis_INTERSTITIAL | Interstitial pancreatitis |
| pancreatitis_INTERSTITIAL_all | Interstitial pancreatitis (all mentions counted) (all notes) |
| pancreatitis_INTERSTITIAL_is_r | Interstitial pancreatitis (number of encounters with mentions) (radiology/imaging only) |
| pancreatitis_INTERSTITIAL_all_is_r | Interstitial pancreatitis (all mentions counted) (radiology/imaging only) |
| panc_with_competing_dx | Pancreatitis with a competing diagnosis |
| panc_without_competing_dx | Pancreatitis without a competing diagnosis |
| panc_with_pain | Pancreatitis with any pain |
| panc_with_radiating_to_back_pain | Pancreatitis with pain described as radiating to the back |
| panc_with_abdominal_pain | Pancreatitis with abdominal pain |

| | |
|---|---|
| acute_panc_without_competing_dx | Acute pancreatitis without a competing diagnosis |
| acute_panc_with_competing_dx | Acute pancreatitis with a competing diagnosis |
| panc_with_sudden_onset_pain | Pancreatitis with sudden onset pain |
| acute_panc_with_sudden_onset_pain | Acute pancreatitis with sudden onset pain |
| acute_panc_imaging | Acute pancreatitis in imaging |
| panc_imaging | Pancreatitis in imaging |
| panc_with_nausea | Pancreatitis with nausea |
| panc_with_necrosis | Pancreatitis with necrosis |
| panc_with_fluid | Pancreatitis with fluid |
| panc_with_pseudocyst | Pancreatitis with pseudocyst |
| panc_with_recency | Pancreatitis with recency mention |
| acute_panc_consistent | Consistent with acute pancreatitis mention |
| panc_consistent | Consistent with pancreatitis mention |
| necrosis_in_imaging | Necrosis in imaging |
| fluid_in_imaging | Fluid in imaging |
| pseudocyst_in_imaging | Pseudocyst in imaging |
| total_text_length | Total length of clinical text processed for this STUDYID measured in characters. |

### Appendix C: Covariates Selected for Custom Modeling

As described in the Methods section of the manuscript, a dimension reduction step was performed to exclude covariates with low variation that were uncorrelated with the outcome. The final data made available to the Super Learner contained the following 220 covariates (59 structured, 161 NLP-derived). Structured covariates are shaded.

| | | |
|---|---|---|
| AGE | I.LIP14 | K85_90 |
| K85_1 | K85_9 | K85_10 |
| FEMALE | ENCTYPE | PRINCIPAL |
| UNSPEC | BILIARY | ETOH |
| LIP14ULN | LIP14X3 | LIP14X10 |
| ABDIMG | GROUP1 | GROUP2 |
| GROUP3 | UPCLASS_EDIP | ENRLYEARS |
| HISPANIC | RACE_UNK | RACE_ASIAN |
| RACE_AA | RACE_WHITE | SMOKE_FORMER_SELF_365 |
| SMOKE_CURR_SELF_365 | ALC_SD | ALC_CS |
| ALC_PY | CHR_PANCR_SD | CONSTIPATION_SD |
| CONSTIPATION_CS | CONSTIPATION_PY | DIVERTICUL_SD |
| DIVERTICUL_PY | GAL_BIL_SD | GAL_BIL_CS |
| GAL_BIL_PY | GASTRITIS_SD | GASTRITIS_CS |
| GASTRITIS_PY | GASTRO_PY | GERD_SD |
| GERD_CS | GERD_PY | HEPATITIS_SD |
| HEPATITIS_CS | HEPATITIS_PY | HYPERTRIG_SD |
| ILEUS_SD | MYOCARD_ISCH_PY | NEPHROLITH_SD |
| NEPHROLITH_CS | NEPHROLITH_PY | ABDIMG_CT_14 |
| ABDIMG_MR_14 | APLAB_MAX_GT3_14BEF_14AFT | PAIN |
| PAIN_ALL | PAIN_IS_R | CDX |
| CDX_ALL | CDX_IS_R | CDX_ALL_IS_R |
| PANCREATITIS | PANCREATITIS_ALL | PANCREATITIS_IS_R |
| NAUSEA | NAUSEA_ALL | NAUSEA_IS_R |
| NECROSIS | NECROSIS_ALL | NECROSIS_IS_R |
| FLUID | FLUID_ALL | FLUID_IS_R |
| PSCYST | PSCYST_IS_R | PAIN_EPIGASTRIC |
| PAIN_EPIGASTRIC_ALL | PAIN_EPIGASTRIC_IS_R | PAIN_CHEST |
| PAIN_CHEST_ALL | PAIN_CHEST_IS_R | CDX_NEGATIVE |
| CDX_NEGATIVE_ALL | CDX_NEGATIVE_IS_R | CDX_GERD |
| CDX_GERD_ALL | CDX_GERD_IS_R | CDX_IBD |
| CDX_IBD_ALL | CDX_IBD_IS_R | PANCREATITIS_ACUTE |
| PANCREATITIS_ACUTE_ALL | PANCREATITIS_ACUTE_IS_R | PANCREATITIS_CONSISTENT |
| PANCREATITIS_CONSISTENT_ALL | PANCREATITIS_CONSISTENT_IS_R | PAIN_RADIATING_TO_BACK |
| PAIN_RADIATING_TO_BACK_ALL | PAIN_RADIATING_TO_BACK_IS_R | NAUSEA_VOMITING |
| NAUSEA_VOMITING_ALL | NAUSEA_VOMITING_IS_R | NAUSEA_NAUSEA |
| NAUSEA_NAUSEA_ALL | NAUSEA_NAUSEA_IS_R | PAIN_ABD_PAIN |
| PAIN_ABD_PAIN_ALL | PAIN_ABD_PAIN_IS_R | CDX_INFLUENZA |
| CDX_INFLUENZA_ALL | CDX_INFLUENZA_IS_R | PANCREATITIS_POSITIVE |
| PANCREATITIS_POSITIVE_ALL | PANCREATITIS_POSITIVE_IS_R | CDX_ASCITES |
| CDX_ASCITES_ALL | CDX_ASCITES_IS_R | CDX_HEPATITIS |
| CDX_HEPATITIS_ALL | CDX_HEPATITIS_IS_R | PAIN_ACUTE |
| PAIN_ACUTE_IS_R | CDX_INFECTIOUS_GE | CDX_PEPTIC_ULCER |
| CDX_PEPTIC_ULCER_ALL | CDX_PEPTIC_ULCER_IS_R | PANCREATITIS_CHRONIC |
| PANCREATITIS_CHRONIC_ALL | PANCREATITIS_CHRONIC_IS_R | CDX_BOWEL_MOVEMENTS |
| CDX_BOWEL_MOVEMENTS_IS_R | CDX_BLOOD_IN_STOOL | CDX_BLOOD_IN_STOOL_ALL |
| CDX_BLOOD_IN_STOOL_IS_R | PAIN_VERY_RECENT | PAIN_VERY_RECENT_IS_R |
| CDX_STROKE | CDX_STROKE_ALL | CDX_STROKE_IS_R |

| | | |
|---|---|---|
| CDX_COLANGITIS | CDX_COLANGITIS_ALL | CDX_COLANGITIS_IS_R |
| CDX_CONSTIPATION | CDX_CONSTIPATION_ALL | CDX_CONSTIPATION_IS_R |
| NECROSIS_ACUTE_COLLECTI | NECROSIS_ACUTE_COLLECTI_ALL | PAIN_RECENT |
| PAIN_CHRONIC | PAIN_CHRONIC_ALL | PAIN_CHRONIC_IS_R |
| CDX_NEPHROLITHIASIS | CDX_NEPHROLITHIASIS_ALL | CDX_NEPHROLITHIASIS_IS_R |
| PANCREATITIS_INFLAMMATI | PANCREATITIS_INFLAMMATI_ALL | PANCREATITIS_INFLAMMATI_IS_R |
| PANCREATITIS_PERI_INFLA | PANCREATITIS_PERI_INFLA_ALL | PANCREATITIS_PERI_INFLA_IS_R |
| PAIN_SUDDEN_ONSET | CDX_DIVERTICULOSIS | CDX_DIVERTICULOSIS_ALL |
| CDX_DIVERTICULOSIS_IS_R | PAIN_UNKNOWN_DURATION | PAIN_UNKNOWN_DURATION_IS_R |
| CDX_CHRONIC_PAIN_ALL | PSCYST_PANCREATIC_IS_R | PSCYST_POSITIVE |
| PSCYST_POSITIVE_ALL | CDX_GALL_BLADDER_DISEAS | CDX_GALL_BLADDER_DISEAS_ALL |
| CDX_GALL_BLADDER_DISEAS_IS_R | CDX_INTESTINAL_OBSTRUCT | CDX_INTESTINAL_OBSTRUCT_ALL |
| CDX_INTESTINAL_OBSTRUCT_IS_R | CDX_MESENTERIC_ISCHEMIA | CDX_MESENTERIC_ISCHEMIA_ALL |
| CDX_ILEUS | CDX_ILEUS_ALL | CDX_ILEUS_IS_R |
| PAIN_WORSENING | CDX_MYOCARDIAL_ISCHEMIA | CDX_MYOCARDIAL_ISCHEMIA_ALL |
| CDX_APPENDICITIS | CDX_APPENDICITIS_ALL | CDX_APPENDICITIS_IS_R |
| CDX_BLOOD_IN_VOMIT | CDX_DKA | CDX_DKA_ALL |
| NECROSIS_POSITIVE_IS_R | CDX_FOOD_POISONING | CDX_ACUTE_APPENDICITIS |
| PAIN_LONG_AGO | CDX_GASTRODUODENITIS | CDX_ESOPHAGITIS |
| CDX_ESOPHAGITIS_ALL | CDX_ESOPHAGITIS_IS_R | PANCREATITIS_INTERSTITI |
| PANC_WITH_COMPETING_DX | PANC_WITHOUT_COMPETING_DX | PANC_WITH_PAIN |
| PANC_WITH_RADIATING_TO_BACK_PAIN | PANC_WITH_ABDOMINAL_PAIN | APANC_WITHOUT_COMPETING_DX |
| APANC_WITH_SUDDEN_ONSET_PAIN | APANC_IMAGING | PANC_IMAGING |
| PANC_WITH_NAUSEA | PANC_WITH_NECROSIS | PANC_WITH_RECENCY |
| NECROSIS_IN_IMAGING | FLUID_IN_IMAGING | PSEUDOCYST_IN_IMAGING |
| TOTAL_TEXT_LENGTH | | |

### Appendix D: AFEP-Derived NLP Dictionary for Automated Modeling

We used the Automated Feature Extraction for Phenotyping (AFEP) approach to identify the following 167 medical terms from clinical knowledge base articles for inclusion in the automated modeling dictionary.

| | | |
|---|---|---|
| 1 (finding) | estrogens | Pancreatitis, Acute |
| 3 times | ethanol | Pancreatitis, Alcoholic |
| Abdomen distended | Facilitated oscillatory release technique | Pancreatitis, Chronic |
| Abdominal Pain | Fatty acid glycerol esters | Parenteral Nutrition, Total |
| Abdominal tenderness | Fever | Pathogenesis |
| Abscess | Fluid resuscitation | pentamidine |
| Active brand of pseudoephedrine-triprolidine | Food allergenic extracts | Pharmaceutical Preparations |
| Adverse Event Associated with Vascular | Functional disorder | Pharmaceutical Solutions |
| Aggressive behavior | furosemide | Physical findings |
| Alcohol consumption | Gallstone pancreatitis | Pleural effusion disorder |
| All of the Time | Gastrointestinal System Finding | Possible |
| Amylase measurement | Hematocrit procedure | Present |
| Amylase measurement, serum (procedure) | Hemorrhage | Pressure- physical agent |
| Analgesics | High Level | Probable diagnosis |
| Antibiotics | Hypercalcemia | Problem |
| APACHE II score | Hyperparathyroidism | Process |
| Aspiration-action | Hypersensitivity | Prophylactic treatment |
| at admission | Hypertriglyceridemia | Proteins |
| azathioprine | Hypocalcemia | Radiating Chest Pain in Abdomen |
| Bacterial Count Measurement | Hypotension | Rather |
| Base | Icterus | Related personal status |
| Biliary Colic | imaging studies | Release (procedure) |
| biomedical tube device | Indicators | Respiratory Distress Syndrome, Adult |
| Blood glucose measurement | Infected | Respiratory System Finding |
| Blood Urea Nitrogen | Infection | risk factors |
| Calcium measurement | Inflammation | Scientific Study |
| Cell Count | Interventional procedure | Severe (severity modifier) |
| chest pain radiating to back | Intestinal Obstruction | Severe disease |
| Cholangiopancreatography, Magnetic Resonance | Invasive Lesion | Severe Extremity Pain |
| Cholecystectomy procedure | Ischemia | Shock |

| | | |
|---|---|---|
| Cholelithiasis | Karnofsky Performance Status 100 | Signs and Symptoms |
| combination - answer to question | Kidney Failure | Single organ dysfunction |
| Communicable Diseases | Kidney Failure, Acute | Skin appearance normal (finding) |
| Complete Blood Count | Laboratory Procedures | Skin necrosis |
| Complication | Laser-Induced Fluorescence Endoscopy | Sulfonamide Anti-Infective Agents |
| Congenital Abnormality | lipase | Symptoms |
| Cullen's sign | Local disease | Systemic Inflammatory Response Syndrome |
| Cystic Fibrosis | Low Level | Systolic blood pressure measurement |
| Cytomegalovirus Infections | Magnetic Resonance Imaging | Tachycardia |
| Data call receiving device | Mass of body structure | Target Lesion Identification |
| Death (finding) | mercaptopurine | Therapeutic procedure |
| Diagnosis | Moderately Severe Hallucination | Tissue damage |
| Diagnostic Imaging | Mumps | Toxic effect |
| Digestive Enzymes | Nausea | Traumatic injury |
| Discharge, body substance | Necrosis of pancreas | Triglycerides |
| Disease | Nutritional Support | trypsin |
| Disorder of small intestine | Obesity | Trypsinogen |
| Drainage procedure | Observation of attack | Ultrasonography |
| Drug Delivery Systems | Operative Surgical Procedures | Unmarried |
| Easy | Pain | Upper abdominal pain |
| Edema | Pain management | Usually |
| Endoscopic Retrograde Cholangiopancreatography | Pancreas divisum | Well Marginated Nodule |
| Endoscopic Ultrasound | Pancreatic Diseases | X-Ray Computed Tomography |
| Enzymes | Pancreatic enzyme | |
| Epidermal cGVHD Score 0 | Pancreatic Pseudocyst | |
| Epidermal cGVHD Score 2 | Pancreatitis | |
| Epidermal cGVHD Score 4 | Pancreatitis Necrotizing | |

### Appendix E: Performance of Custom Modelling Approaches

We used the Super Learner R package to consider multiple machine learning algorithms for prediction. The Super Learner library consisted of eight parametric and machine learning algorithms [logistic regression (GLM), elastic net (Enet), two variants of gradient boosting (XGBoost), two variants of bayesian additive regression trees (BART), and two neural net architectures (NNET)] coupled with three covariate retention strategies [Retain all, partitioning around medoids (PAM), retain those with non-zero coefficients based on their conditional associations with the outcome (Lasso)]. Below is the cross-validation AUC with 95% confidence interval for each algorithm-retention pair using structured covariates alone (Table 1) or structured plus NLP covariates (Table 2).

Table 1. cv-AUC (95%CI) for each algorithm-retention pair using structured covariates alone

| | **Retention Strategy** | | |
|---|---|---|---|
| **Algorithm** | **Retain All** | **PAM** | **Lasso** |
| GLM | 0.85 (0.80, 0.90) | 0.85 (0.80, 0.90) | 0.89 (0.84, 0.94) |
| Enet | 0.89 (0.84, 0.94) | 0.89 (0.84, 0.94) | 0.89 (0.84, 0.94) |
| XGBoost-1 | 0.89 (0.84, 0.94) | 0.89 (0.84, 0.94) | 0.89 (0.85, 0.95) |
| XGBoost-2 | 0.90 (0.85, 0.95) | 0.90 (0.85, 0.95) | 0.89 (0.84, 0.94) |
| BART-1 | 0.89 0.84, 0.94) | 0.89(0.84, 0.94) | 0.90 (0.85, 0.95) |
| BART-2 | 0.90 (0.85, 0.95) | 0.90 (0.85, 0.95) | 0.90 (0.85, 0.95) |
| NNET-1 | 0.80 (0.75, 0.84) | 0.82 (0.78, 0.87) | 0.90 (0.82, 0.92) |
| NNET-2 | 0.81 (0.76, 0.85) | 0.82 (0.77, 0.86) | 0.90 (0.85, 0.95) |
| SL | 0.89 (0.84, 0.94) | | |

Table 2. cv-AUC (95%CI) for each algorithm-retention pair using structured plus NLP covariates

| | **Retention Strategy** | | |
|---|---|---|---|
| **Algorithm** | **Retain All** | **PAM** | **Lasso** |
| GLM | 0.72 (0.68, 0.76) | 0.72 (0.68, 0.77) | 0.90 (0.85, 0.95) |
| Enet | 0.91 (0.85, 0.96) | 0.83 (0.78, 0.88) | 0.91 (0.86, 0.97) |
| XGBoost-1 | 0.92 (0.87, 0.97) | 0.83 (0.79, 0.88) | 0.92 (0.87, 0.97) |
| XGBoost-2 | 0.92 (0.87, 0.97) | 0.84 (0.79, 0.89) | 0.92 (0.87, 0.97) |
| BART-1 | 0.93 (0.88, 0.98) | 0.85 (0.80, 0.89) | 0.92 (0.87, 0.98) |
| BART-2 | 0.92 (0.87, 0.98) | 0.85 (0.80, 0.90) | 0.93 (0.88, 0.98) |
| NNET-1 | | | 0.90 (0.85, 0.95) |
| NNET-2 | | | |
| SL | 0.93 (0.88, 0.98) | | |

### Appendix F: Retained Covariates in Custom Modelling (Enet-Lasso)

Below are the logistic regression coefficients for retained covariates in the custom modelling Enet-Lasso algorithm-retainer pair using structured covariates alone (Table 1) and structured plus NLP covariates (Table 2). Variable importance rankings were defined as the marginal mean difference in predicted probability associated with a 1-unit change in a binary covariate or a 1-standard-deviation change in a nonbinary covariate. Variable importance rankings are presented below for retained covariates in the custom modelling Enet-Lasso algorithm-retainer pair using structured covariates alone (Table 3) and structured plus NLP covariates (Table 4).

#### Table 1. Logistic regression coefficients for structured covariates alone

| Covariate | Coefficient |
|---|---|
| intercept | -2.402299 |
| PRINCIPAL | 2.2289255 |
| LIP14X3 | 3.4409809 |
| GROUP1 | -0.3549629 |
| UPCLASS_EDIP | 1.7225699 |
| DIVERTICUL_SD | 1.3099461 |
| DIVERTICUL_PY | 0.7647781 |
| GAL_BIL_PY | 1.1387617 |
| GERD_PY | 0.5005527 |
| HYPERTRIG_SD | 5.0376709 |
| ABDIMG_CT_14 | 0.5728201 |
| APLAB_MAX_GT3_14BEF_14AFT | 1.3555024 |

#### Table 2. Logistic regression coefficients for structured plus NLP covariates

| Covariate | Coefficient |
|---|---|
| intercept | -2.583383 |
| PRINCIPAL | 1.579435e+00 |
| LIP14X3 | 3.886071e+00 |
| GROUP1 | 3.230652e-01 |
| HYPERTRIG_SD | 4.114196e+00 |
| APLAB_MAX_GT3_14BEF_14AFT | 8.600643e-01 |
| PANCREATITIS_ACUTE_IS_R | 1.727960e+05 |
| PANCREATITIS_CONSISTENT_IS_R | 2.474539e+04 |
| PANCREATITIS_INFLAMMATI_IS_R | 7.840011e+04 |
| PANCREATITIS_PERI_INFLA | 1.012931e+05 |
| CDX_GALL_BLADDER_DISEAS_ALL | 6.086987e+04 |
| CDX_APPENDICITIS_IS_R | -6.024004e+04 |
| PANCREATITIS_INTERSTITI | 1.538020e+05 |

Table 3. Variable importance rankings for structured covariates alone

| Covariate | Marginal RD |
|---|---|
| LIP14X3 | 0.538 |
| HYPERTRIG_SD | 0.384 |
| PRINCIPAL | 0.236 |
| UPCLASS_EDIP | 0.173 |
| APLAB_MAX_GT3_14BEF_14AFT | 0.150 |
| DIVERTICUL_SD | 0.128 |
| GAL_BIL_PY | 0.110 |
| DIVERTICUL_PY | 0.073 |
| ABDIMG_CT_14 | 0.056 |
| GERD_PY | 0.048 |
| GROUP1 | -0.033 |

Table 4. Variable importance rankings for structured plus NLP covariates

| Enet - Lasso | |
|---|---|
| **Covariate** | **Marginal RD** |
| LIP14X3 | 0.569 |
| HYPERTRIG_SD | 0.340 |
| PRINCIPAL | 0.132 |
| PANCREATITIS_ACUTE_IS_R | -0.089 |
| APLAB_MAX_GT3_14BEF_14AFT | 0.072 |
| PANCREATITIS_INTERSTITI | -0.048 |
| PANCREATITIS_PERI_INFLA | -0.040 |
| PANCREATITIS_INFLAMMATI_IS_R | -0.039 |
| PANCREATITIS_CONSISTENT_IS_R | -0.031 |
| CDX_GALL_BLADDER_DISEAS_ALL | -0.029 |
| GROUP1 | 0.025 |
| CDX_APPENDICITIS_IS_R | 0.012 |
| PANCREATITIS_IS_R | -0.001 |
